# iSPHYNCS: Unsupervised clustering in questionnaires and metadata reveals distinct subtypes in the narcolepsy borderland

**DOI:** 10.1101/2025.03.25.25324595

**Authors:** Rafael Morand, Livia Fregolente, Julia van der Meer, Elena S. Wenz, Annina Helmy, Lorenzo Brigato, Jan D. Warncke, Kseniia Zub, Ramin Khatami, Zhongxing Zhang, Sigrid von Manitius, Silvia Miano, Jens Acker, Mathias Strub, Ulf Kallweit, Gert Jan Lammers, Athina Tzovara, Claudio L.A. Bassetti, Stavroula Mougiakakou, Markus H. Schmidt

## Abstract

**Objective:** The international Swiss Primary Hypersomnolence and Narcolepsy Cohort Study (iSPHYNCS) is a multicenter study aimed at identifying novel biomarkers for central disorders of hypersomnolence (CDH). We analyzed questionnaires and metadata to uncover distinct clusters of participants and explore phenotypic variability within CDH.

**Methods:** Data were collected from 227 patients with CDH and 33 healthy controls. Participants completed validated clinical questionnaires and study-specific questions addressing CDH-related symptoms such as excessive daytime sleepiness, fatigue, cataplexy, disrupted sleep, and sleep paralysis. Demographic metadata (age, gender, BMI) were included. After excluding participants with missing over 30% of data (n = 40), missing values were imputed using a multiple random forest algorithm. A robust clustering pipeline was employed: (1) random sampling of 60% of the dataset, (2) dimensionality reduction via UMAP, (3) K-means clustering, and (4) consensus clustering across 500 iterations. Post hoc analysis was performed to identify biomarkers in data not used for clustering.

**Results:** We identified four distinct clusters. One predominantly comprised healthy controls, while another primarily contained individuals with narcolepsy type 1 (NT1). Two clusters represented predominantly the narcolepsy borderland group (NBL), with one distinctly characterized by higher symptom severity and psychiatric comorbidities.

**Conclusions:** The clustering pipeline produced reproducible results, with the NT1 and healthy control clusters serving as internal validation. The differentiation between the two NBL clusters aligns with prior studies, suggesting a possible NBL subtype marked by increased fatigue and psychiatric comorbidities. These findings emphasize the phenotypic heterogeneity of CDH and the potential for cluster-based approaches in management.

**Statement of Significance:** Central disorders of hypersomnolence (CDH) are complex and heterogeneous, challenging conventional diagnostic frameworks. This study employed an innovative clustering pipeline to identify four distinct phenotypic clusters within a large, multicenter cohort. The identification of a subtype in the narcolepsy borderland group with a high psychiatric burden adds critical insights into the variability of CDH and underscores the need for personalized diagnostic and therapeutic strategies. The internal validation of clusters through healthy controls and NT1 phenotypes enhances confidence in the pipeline’s robustness. Furthermore, the study highlights the potential for integrating clinical data to define disease severity and developing therapeutic strategies. These findings pave the way for novel approaches to understanding and managing CDH.

## Introduction

Central disorders of hypersomnolence (CDH) represent a group of diseases characterized by an overwhelming and irrepressible need for sleep that impacts daily functions. These disorders are currently classified according to the International Classification of Sleep Disorders, third edition (ICSD-3)^1^. Nevertheless, due to often overlapping and ambiguous diagnostic criteria, there have been numerous recent efforts attempting to reassess and refine the current classification system^2,3^. While experts agree that narcolepsy type 1 (NT1) can be reliably differentiated by the presence of cataplexy and absence or low levels of hypocretin in the cerebrospinal fluid^4^, other CDH disorders from the so-called narcolepsy borderland (NBL), such as narcolepsy type 2 and idiopathic hypersomnia, rely heavily on subjective assessments and lack pathophysiological specificity^5^.

Recent advancements in data science, particularly machine learning techniques, have introduced new possibilities for phenotyping sleep-wake disorders based on clinical and biological data^6,7^. Clustering algorithms, for instance, are designed to group patients into phenotypes that exhibit minimal intra-group variability and well-defined inter-group differences^8^. In the context of CDH, unsupervised clustering methods have shown promise in reclassifying patients into phenotypes that may be more clinically meaningful than traditional observations^9,10^. More specifically, identified clusters distinguished patients within the NBL based on the presence or absence of sleep drunkenness^9^, as well as demographical and electrophysiological markers^10^.

A further study employing unsupervised clustering approaches in patients diagnosed with hypersomnolence according to the fifth edition of Diagnostic and Statistical Manual of Mental Disorders (DSM-5) identified clusters differentiated by variables such as the presence of depression and other clinical features^11^. These findings underscore the importance of considering psychiatric comorbidities, which have been shown to exacerbate the impact of CDH and contribute to a vicious cycle of fatigue, sleepiness and reduced quality of life^12^.

While unsupervised learning techniques have provided promise in refining diagnostic categories for CDH, it remains unclear whether these phenotypes are generalizable and applicable in other populations. Moreover, previous observations have relied on the presence of electrophysiological and biological data without a control group for comparison. Therefore, the aim of this study was to apply unsupervised clustering techniques to reclassify patients with CDH based on a comprehensive set of clinical data, including a healthy control group. To achieve this, we utilized the well-characterized international Swiss Primary Hypersomnolence and Narcolepsy Cohort Study (iSPHYNCS)^3^, a multicenter prospective cohort study involving collaboration across ten centers in three European countries. We hypothesized that by using data from questionnaires and metadata, unsupervised clustering would a) result in a clear separation of healthy controls from patient groups, b) result in a clear separation of individuals diagnosed with NT1 from those without cataplexy, and c) group individuals from the NBL into multiple clusters according to the perceived severity of symptoms. We are confident that the findings from our cluster analysis will provide a refined foundation for future therapeutic frameworks of the NBL.

## Methods

### Study Design and Setting

This study was conducted across ten sleep centers located in three European countries—Switzerland, Germany, and the Netherlands—as part of the prospective international Swiss Primary Hypersomnolence and Narcolepsy Cohort Study (iSPHYNCS). The iSPHYNCS is a multicenter study aiming at investigating the phenotypes and clinical characteristics of individuals with central disorders of hypersomnolence. The complete study protocol has been previously published^3^. This analysis uses data from participants enrolled at the following sleep clinics and study sites (listed in alphabetical order): Bad Zurzach (Switzerland); Barmelweid (Switzerland); Basel (Switzerland); Bern (Switzerland); Heemstede (the Nederlands); Lugano (Switzerland); St. Gallen (Switzerland); and Witten (Germany).

### Participants

We included individuals aged 16–70 years who presented to the participating sleep centers with subjective complaints of excessive daytime sleepiness (EDS) and/or hypersomnolence. Patients included in this analysis were consecutively recruited into the study between September 2020 and October 2024 and needed to have completed at least 70% of the study’s questionnaires. Eligible participants were fluent speakers of German, French, Dutch, or Italian and provided informed written consent prior to participation. Exclusion criteria included the presence of other sleep disorders, neurological conditions, or systemic diseases that could more likely explain their EDS or hypersomnolence. Healthy controls were recruited from local advertising and excluded if showing signs of EDS or significant SDB.

Participants were provisionally categorized based on the International Classification of Sleep Disorders – Third Edition (ICSD-3)^13^ into three groups: NT1, healthy controls (HC), and the broader narcolepsy borderland (NBL). The NBL group encompassed diagnoses such as narcolepsy type 2, idiopathic hypersomnia, insufficient sleep syndrome, hypersomnia associated with a psychiatric disorder. We also included patients who did not meet criteria for a specific CDH; these patients were classified as excessive daytime sleepiness not otherwise specified.

Reviewed diagnoses were established by a panel of at least two clinician experts using ICSD-3 criteria. For NT1, the diagnosis was also considered as reviewed if available cerebrospinal fluid hypocretin-1 levels were below 110 pg/mL^4^. In the other cases, provisional diagnoses assessed by the treating physician were considered for analysis as unreviewed.

### Data Collection

Data were collected prospectively using standardized questionnaires and a systematic interview. In December 2022, the study expanded to include international collaborators, transitioning from SPHYNCS to iSPHYNCS, and the range of assessment tools was adjusted. For instance, the Mini International Neuropsychiatric Interview (MINI)^14^ and the Beck depression inventory-II^15^ were replaced with the hospital anxiety and depression scale (HADS)^16^.

This analysis includes only responses obtained at inclusion and for questions completed by at least 60% of the cohort. The nine questionnaires included in the clustering pipeline, and for which licenses have been obtained, are: Epworth sleepiness scale (ESS)^17^, fatigue severity scale (FSS)^18^, narcolepsy severity scale (NSS)^19^, Swiss narcolepsy scale(SNS)^20^, Pittsburgh sleep quality index (PSQI)^21^, idiopathic hypersomnia severity scale (IHSS)^22^, sleep inertia questionnaire (subset of SIQ)^23^, 36-item short form survey instrument (subset of SF36)^24^, and functional outcome of sleep questionnaire (FOSQ)^25^. These instruments and other metadata questions captured a broad range of subjective clinical variables, including sleep quality, hypersomnolence severity, and functional impairment. An extensive description of the questionnaires is provided in the supplementary material (S1).

### Unsupervised Clustering

We employed an unsupervised clustering pipeline to identify distinct phenotypic subgroups within the study cohort (Fig. 1). A detailed description of this approach is provided in the supplementary material (S2).

**Figure 1.**
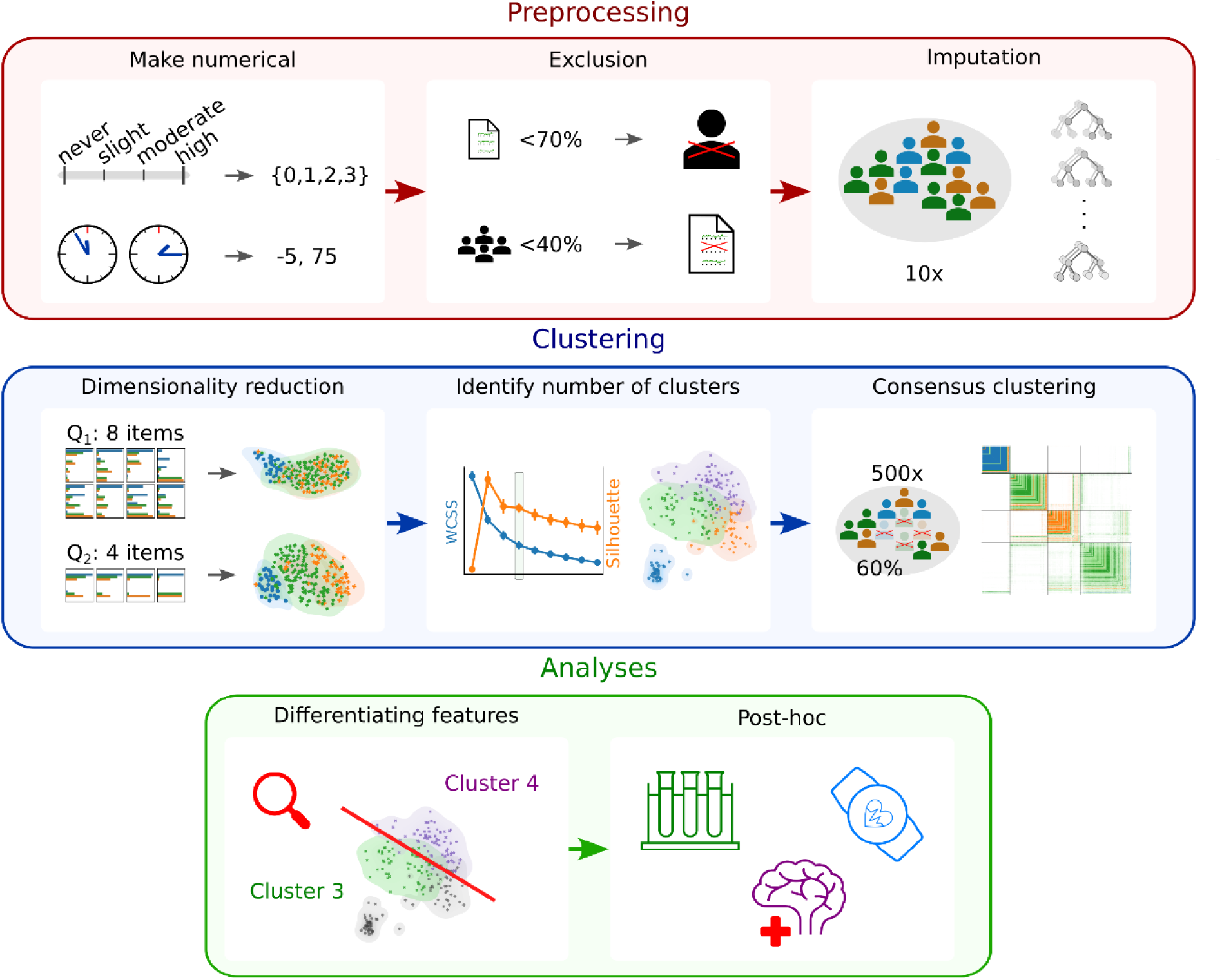
Schematic of the preprocessing and clustering pipeline, and the subsequent analyses.

For the preprocessing, we encoded categorical data to be interpreted as numerical values, and timestamps were transformed into minutes relative to midnight. Then, we excluded individuals if they had answered less than 70% of the questions. In addition, we excluded variables if they were answered by less than 40% of the study participants. For imputation, we performed multiple imputation with 10 iterations using a random forest imputer^1,10^. For normalization, we scaled each variable to a range from −1 to 1 across all cohorts.

Reducing the dimensions for all questionnaires to a common fixed size is beneficial as it gives equal weights to all questionnaires in the clustering algorithm. Hence, we reduced the dimensionality using UMAP^26^ on each questionnaire individually to compress the N-dimensional questionnaires into 2 dimensions, where N denotes the number of items per questionnaire (supplementary material S3).

To cluster the compressed data, we initially identified the number of clusters (= *k*) by using the full cohort and applying the K-means algorithm with 50 different initial conditions (five random seeds for ten imputations). Using this prior knowledge, we then performed consensus clustering. For this, we repeatedly used the K-means algorithm on 500 randomly drawn subsets of the data (50 random seeds for ten imputations) that included only 60% of the individuals and searched for *k* clusters. We set the conclusive cluster based on the consensus across repetitions (i.e., individuals who repeatedly clustered together received the same conclusive cluster label). Individuals with extraordinarily low consensus with their clusters were excluded from further analyses (n = 16).

### Identification of most differentiating features

To explain the differences between the clusters, we performed statistical tests on the data used in the clustering process. This analysis focused on the differences between the clusters that mainly contained individuals from the NBL since this was the group of interest.

Depending on the type of each variable, we calculated an appropriate effect size and set conservative thresholds to categorize them as either strongly differentiating or not differentiating. For intervals and ratios, we calculated the standardized effect size using Cohen’s d with pooled variance for groups with unequal variance and unequal sample sizes (d ≥ 0.8 and d ≤ 0.2 for strongly diff. and not diff.). For ordinal categories, we calculated the odds ratio using ordinal logistic regression (OR > 4 and OR < 2 for strongly diff. and not diff.; applying the reciprocal). For nominal data with more than two categories, we calculated Carmér’s V using chi-square (V > 0.5 and V < 0.1 for strongly diff. and not diff.). For nominal data with two categories, we calculated the odds ratio using Fisher’s exact test (OR > 10 and OR < 2 for strongly diff. and not diff.; applying the reciprocal).

### Post hoc Analysis

To explore further differences between the identified clusters, we analyzed variables that were not used for clustering. Specifically, we assessed the impact of psychiatric comorbidities, established NT1 biomarkers, and objective sleep measures.

The impact of psychiatric symptoms and comorbidities was assessed using data derived from either the MINI, the HADS, or the BDI^14,16^. These questionnaires regarding psychiatric comorbidities were previously excluded from the clustering because, after the internationalization of the iSPHYNCS, the study participants received only a subset of these questionnaires due to changes in the study protocol. Therefore, if we included them in the clustering pipeline, participants might have clustered based on the questionnaires they received and not on the responses.

Where available, hypocretin-1 levels and human leukocyte antigen (HLA) DQB1*0602 phenotyping were included, as these are established biomarkers for NT1^4^.Regarding objective sleep measures, we compared the mean sleep latency from the multiple sleep latency test (MSLT)^27^ between the clusters. In addition, data from physical activity recordings (Fitbit Inspire HR/2/3) worn by participants over the course of up to one year were analyzed to extract sleep parameters such as sleep duration and bedtime/get-up time. Participants were instructed to wear the Fitbit on the non-dominant arm. These data were acquired using a custom pipeline^28^.

The normality of distributions was assessed using Shapiro-Wilk tests. For normally distributed data, p-values were calculated using Welch’s t-test for unequal variances. For non-normally distributed data, p-values were derived from Mann-Whitney U tests.

## Ethical Approval

The study protocol received ethical approval from the relevant authorities in each participating country, and all participants provided informed consent in accordance with the Declaration of Helsinki. Ethical approval for the study was obtained from local ethics committees (Ethics Committees ID: 2019–00788 in Switzerland, 202/2022 in Germany, and NL84710.058.23 in the Netherlands), and the trial is registered in the international clinical trials registry (ClinicalTrials.gov Identifier: NCT04330963).

## Data Availability

The data underlying this article will be shared on reasonable request to the corresponding author.

## Results

### Population

Originally, 260 participants were included in the iSPHYNCS cohort as of October 2024. Of these, 40 participants were excluded: 39 due to insufficient data and one due to a hypersomnia diagnosis that was ultimately attributed to a systemic disease, which constituted an exclusion criterion. Thus, the final analysis included 220 participants, whose baseline characteristics are summarized in Table 1.

**Table 1.** Baseline demographics.

|  | Missing | Overall | Healthy controls | Narcolepsy Type 1 | Narcolepsy<br>borderland |
| --- | --- | --- | --- | --- | --- |
| **n** |  | 220 | 33 | 51 | 136 |
| **Age, median [Q1,Q3]** | 0 | 26 [21,33] | 28 [24,32] | 26 [21,35] | 26 [21,33] |
| **BMI, median [Q1,Q3]** | 5 | 23.6 [21.1,27.2] | 22.9 [21.3,25.8] | 23.9 [21.9,28.5] | 23.7 [20.7,27.5] |
| **Gender (f), n (%)** | 0 | 155 (70.5) | 20 (60.6) | 30 (58.8) | 105 (77.2) |
| **Work active, n (%)** | 12 | 148 (71.2) | 26 (81.2) | 27 (55.1) | 95 (74.8) |
| **Education, n (%)** | 7 |  |  |  |  |
| *Secondary* |  | 43 (20.2) | 0 (0.0) | 12 (23.5) | 31 (23.8) |
| *Apprenticeship* |  | 73 (34.3) | 6 (18.8) | 17 (33.3) | 50 (38.5) |
| *High school* |  | 36 (16.9) | 8 (25.0) | 9 (17.6) | 19 (14.6) |
| *University* |  | 58 (27.2) | 17 (53.1) | 13 (25.5) | 28 (21.5) |
| *Other* |  | 3 (1.4) | 1 (3.1) | 0 (0.0) | 2 (1.5) |
| **Hypocretin, median [Q1,Q3]** | 133 | 270 [88,320] | *na* | 44 [19.5,88] | 299 [273,333] |
| **HLA:DQB1\*06.02 positive, n (%)** | 73 | 54 (36.7) | 3 (20.0) | 31 (96.9) | 20 (20.0) |
| **Medication, n (%)** | 0 |  |  |  |  |
| *No medication* |  | 116 (52.7) | 33 (100.0) | 13 (25.5) | 70 (51.5) |
| *Antidepressant* |  | 21 (9.5) | 0 (0.0) | 3 (5.9) | 18 (13.2) |
| *Sodium Oxybate* |  | 3 (1.4) | 0 (0.0) | 2 (3.9) | 1 (0.7) |
| *Stimulant* |  | 66 (30.0) | 0 (0.0) | 32 (62.7) | 34 (25.0) |
| *Other* |  | 14 (6.4) | 0 (0.0) | 1 (2.0) | 13 (9.6) |

The median age of participants was 26 years (interquartile range [IQR]: 21–33), and 70.5% were female. The cohort consisted of 51 participants with NT1, 33 HC, and 136 participants in the NBL. Most participants were employed or studying, with 71.2% reporting active engagement in work or education, although this was lower in the NT1 group (55.1%) compared to HCs (81.2%) and the NBL group (74.8%).

### Cluster Analysis

The consensus clustering resulted in four distinct clusters, as illustrated in Figure 2. Below, we describe the characteristics of each cluster in detail.

**Figure 2.**
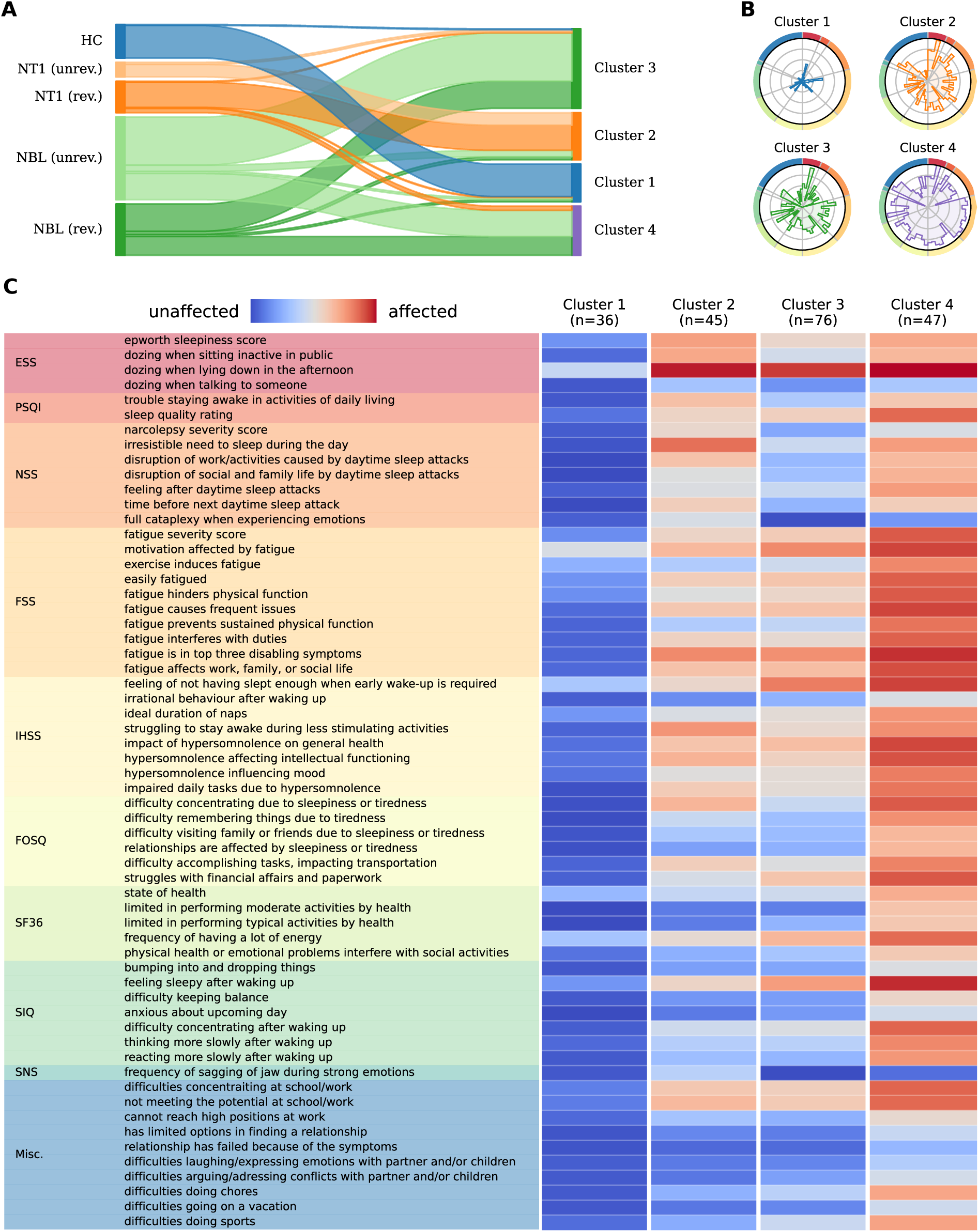
Description of the four identified clusters. A) The flowchart shows how the study cohorts are distributed into the clusters. The patient groups are further divided if reviewed or not of the diagnosis (rev. = reviewed diagnosis, unrev. = unreviewed diagnosis). B) The radar plots highlight how the clusters have unique footprints regarding the perceived affectedness (see C for the color-coded questions). Cluster 1 is virtually unaffected, clusters 2 and 3 are mild phenotypes that differ in the affectedness regarding cataplexy, and cluster 4 reports overall high affectedness. C) The barcode plot provides a description of the most differentiating features. The color code from blue to red corresponds to the magnitude of the radar plots in B. *Abbreviations: HC=Healthy controls, NT1=Narcolepsy Type 1, NBL=Narcolepsy Borderland, ESS=Epworth Sleepiness Scale, PSQI=Pittsburgh Sleep Quality Index, NSS=Narcolepsy Severity Scale, FSS=Fatigue Severity Scale, IHSS=Idiopathic Hypersomnia Severity Scale, FOSQ=Functional Outcome of Sleep Questionnaire, SF36=Short Form-36, SIQ=Sleep Inertia Questionnaire, SNS=Swiss Narcolepsy Scale, Misc.=Miscellaneous*.

### Cluster 1

This cluster (n = 36) included participants who were largely unaffected across all domains analyzed. Nearly all HC (n = 31) fell into this group, with only one excluded due to low consensus, and another being in a different cluster. Both expressed higher levels of anxiety and pain. Additionally, this cluster included four individuals from the NBL and one patient with NT1, reflecting a general absence of significant symptoms.

### Cluster 2

This cluster (n = 45) was primarily characterized by pronounced daytime sleepiness and a higher likelihood of experiencing cataplexy. Participants frequently reported emotion-triggered muscle weakness and an increased propensity to fall asleep during the day. In contrast, compared to cluster 4, these individuals reported lower levels of fatigue and fewer restrictions in daily life. This cluster mainly contains patients categorized as NT1 and ten patients from the NBL.

### Cluster 3

This cluster (n = 76) included participants predominantly from the NBL exhibiting a milder phenotype. Compared to cluster 4, patients in cluster 3 reported fewer restrictions in daily activities, better sleep quality, and an absence of emotion-triggered muscle weakness (Figure 2, table S1). Notably, gender, medication use, questions from the PSQI and morningness-eveningness preferences did not significantly differentiate between this group from cluster 4 (table S2). Three patients with NT1 and one healthy control were assigned to this cluster.

### Cluster 4

This cluster (n = 47) comprised predominantly participants from the NBL with a more severe symptom profile. It also included four individuals with an NT1 diagnosis. Participants in this group exhibited significantly higher levels of fatigue, with both cognitive and physical components of fatigue contributing to increased exhaustibility. Additionally, this cluster was characterized by excessive daytime sleepiness, sleep drunkenness, poor sleep quality, and a pronounced sense of restriction and underachievement in daily activities. Individuals in this group reported substantial functional impairments across multiple domains, including cognition, social interactions, and workplace or academic performance.

Comprehensive tables for all differentiating features are provided in the supplementary material (S4).

### Individuals with unexpected cluster assignment

There were eight individuals with a NT1 diagnosis who were assigned to different clusters than cluster 2, which contained the majority of NT1 patients. In cluster 4, there were four (8.5%) participants with a diagnosis of NT1 (three definite with low hypocretin levels, one probable) who reported enhanced fatigue, sleep drunkenness, and impairment in daily activities.

Three individuals with NT1 diagnoses was assigned to cluster 3 (two definite, one probable), and another with a confirmed NT1 diagnosis and low hypocretin levels was placed in cluster 1. All four individuals were receiving stimulant treatment, suggesting either fluctuating or milder phenotypes influenced by medication effects. Additionally, four participants from the NBL were assigned to cluster 1 due to a less affected phenotype. Notably, only one of these participants was taking medication at the time of assessment.

Furthermore, ten patients from the NBL were assigned to cluster 2, primarily because they reported muscle weaknesses triggered by emotions but did not report the pronounced fatigue observed in cluster 4. There was one healthy control in cluster 3 that reported higher levels of pain and anxiety. Proportionally, no significant center differences were observed among individuals with unexpected cluster assignment.

### Post hoc Analysis

To better characterize the difference between the two borderland clusters, we evaluated the biomarkers HLA:DQB1*0602 and hypocretin-1 levels, as well as the burden of psychiatric symptoms and comorbidities using validated questionnaires (Fig. 3).

**Figure 3.**
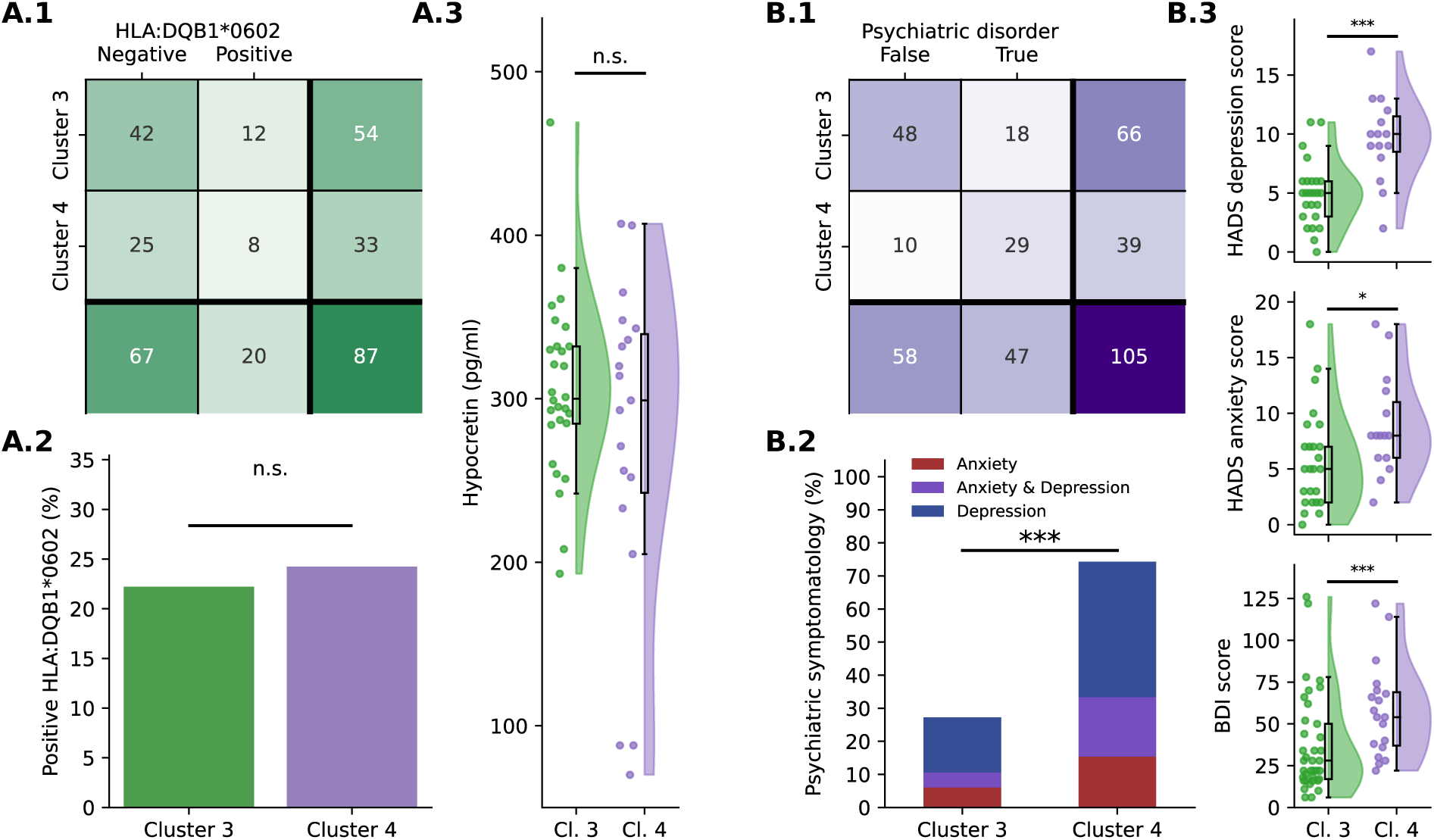
Comparison of NT1 biomarkers and psychiatric disorders between cluster 3 and cluster 4. A) (1) Contingency table of the HLA:DQB1*0602 positivity. The total number of individuals was lower since the analysis was not available for every individual (43/61 and 46/71). (2) The barplot with the proportion of HLA:DQB1*0602 positive individuals shows that there is no significant difference between the clusters. (3) The hypocretin levels were not significantly different between the clusters. B) (1) Contingency table showing the presence of psychiatric symptomatology based on the HADS and MINI questionnaires. The total number of individuals was lower since not every individual provided answers to the questionnaires (54/61 and 60/71). (2) The barplot shows that cluster 4 had a significantly higher proportion of individuals with a psychiatric symptomatology. (3) Cluster 4 scored higher in the HADS questionnaire for depression and also in the BDI compared to cluster 3.

There was no significant difference regarding HLA:DQB1*0602 positivity (22.2% vs. 24.2%, p=1.0). Moreover, there was no significant difference between the groups regarding the hypocretin-1 levels (305 ± 54 vs. 275 ± 101, p=0.625). Most participants (105/123, 85.4%) had either results from the MINI, BDI or the HADS score to calculate the burden of psychiatric symptomatology. Cluster 4 had a significantly higher prevalence of either anxiety or depression than cluster 3 (74.4% vs 27.3%, p < 0.001), with a higher mean HADS depression score (9.60 ± 3.60 vs. 4.96 ± 2.75, p < 0.001), HADS anxiety score (8.87 ± 4.50 vs. 5.84 ± 4.45, p = 0.030), and BDI score (61.7 ± 32.0 vs. 34.2 ± 27.9, p < 0.001).

There was no significant difference between cluster 3 and cluster 4 in the REM propensity (5.3% vs. 8.5%, p=0.480) nor in the mean sleep latency of the MSLT (9.2 ± 4.2 vs. 9.5 ± 5.4, p=0.919), as shown in Figure 4.A. Although not statistically significant, it seems like cluster 4 demonstrates a higher variability in sleep duration (108 ± 30 minutes vs. 91 ± 25 minutes, p = 0.078) and bedtime (105 ± 51 minutes vs. 85 ± 43 minutes, p = 0.300), indicating poorer sleep hygiene (Figure 4.B).

**Figure 4.**
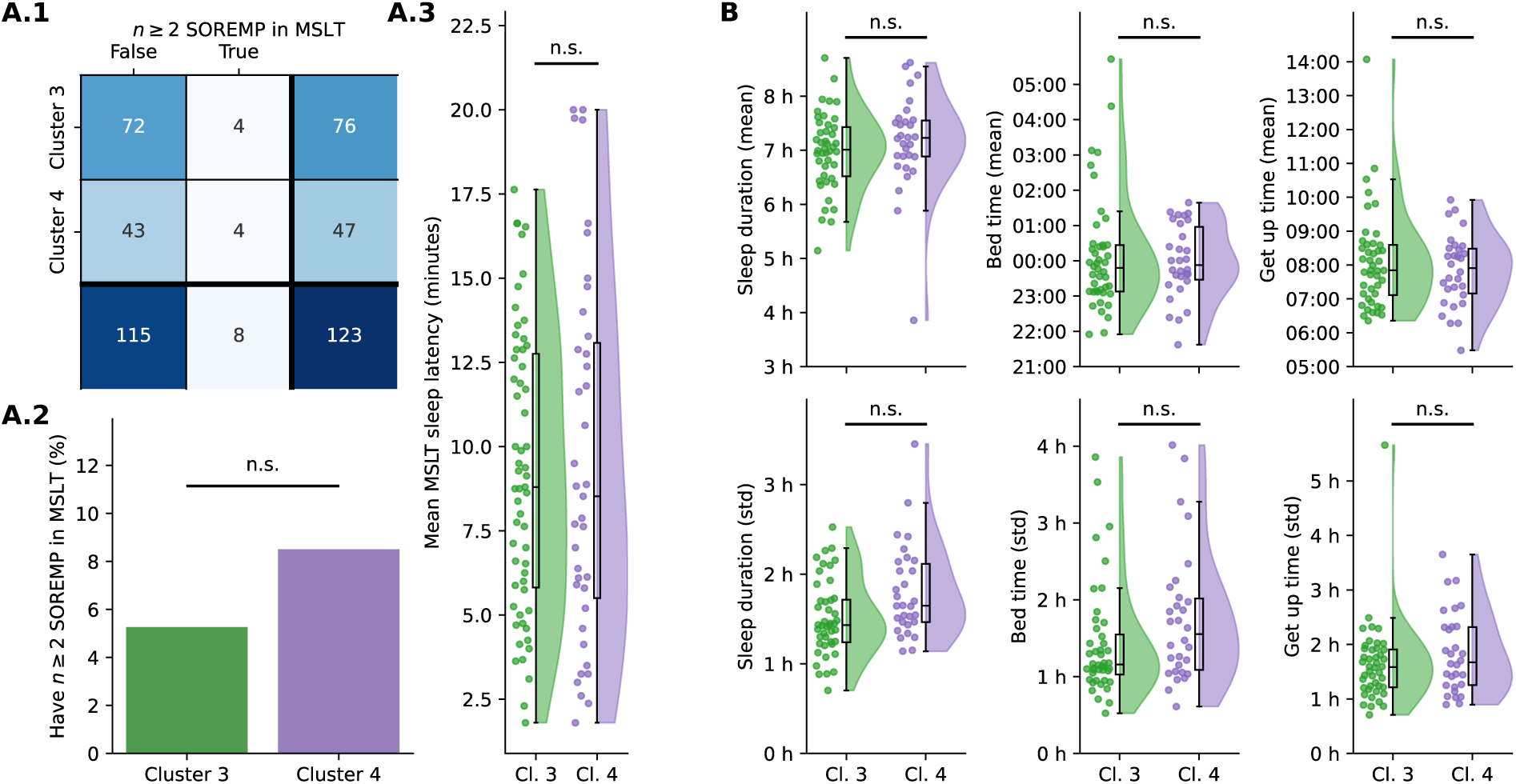
Objective sleep measures. A) Sleep parameters from MSLT. There were no significant differences between the clusters regarding the number of SOREMPs and the mean MSLT sleep latency. B) Sleep parameters from long-term physical activity recordings. There were no significant differences, although it seems like cluster 4 demonstrates a higher variability in sleep duration and bedtime, indicating poorer sleep hygiene..

## Discussion

This study aimed to examine the clinical heterogeneity of central disorders of hypersomnolence (CDH) by employing unsupervised clustering methods to identify distinct phenotypic subgroups. It is the first study to develop a computational pipeline powerful enough to differentiate between groups by utilizing only readily available data from questionnaires. The findings highlight the diverse symptom profiles among individuals with narcolepsy and its borderland, including, for the first time, healthy controls, emphasizing the utility of clustering in refining diagnostic and therapeutic approaches.

The identification of clusters underscores the clinical variability within the CDH spectrum. Cluster 4 represents a subgroup with questionnaire data pointing towards severe functional impairments and morbidity, as evidenced by high levels of cognitive and physical fatigue, sleep drunkenness and low sleep quality. Daily activities, including work, school and social interactions, as well as cognitive functions (e.g., remembering, concentrating), seem significantly affected. Cluster 4 is the only cluster also showing increased physical exhaustibility (e.g., FSS: “exercise induces fatigue”, “fatigue prevents sustained physical function”). These features, although described in CDH^29^, are more commonly observed in affective disorders and other fatigue-related conditions (e.g., post-infectious fatigue, chronic fatigue syndrome). Notably, this cluster included mainly individuals from the narcolepsy borderland but also a few definite NT1 (low hypocretin). These findings suggest that severe functional impairments may overshadow specific biomarkers, including hypocretin deficiency. Cluster 4 may represent patients characterized by significant morbidity likely requiring more intensive clinical management^30,31^. Affective disorders should be optimally treated, if necessary pharmaceutically^30,32^. The pattern and severity of complaints (e.g., fatigue, exhaustibility) might also require some additional and tailored therapeutic strategies, like psychotherapy, physiotherapy, and pacing strategies.

Cluster 3, in contrast, represents a milder phenotype, with less impact on daily functioning, suggesting a distinct pathophysiological or psychosocial trajectory compared to cluster 4. Supporting those first findings, the post-hoc analyses revealed significant differences in psychiatric comorbidity between the two borderland clusters. Cluster 4 had a notably higher prevalence of anxiety or depression compared to Cluster 3 (74.4% vs 27.3%), confirmed by elevated scores on validated psychiatric scales such as the HADS and BDI. This aligns with previous literature suggesting a strong association and probably bi-directional relationship between hypersomnolence and mood disorders, particularly in individuals with greater functional impairment^12,33^.

Cluster 2 was characterized by pronounced cataplexy, excessive daytime sleepiness, and increased susceptibility to unintentional daytime sleep episodes, closely aligning with the clinical presentation of narcolepsy type 1^4^. This cluster reinforces the diagnostic relevance of cataplexy as a key distinguishing feature among CDH subgroups^34^. However, the assignment of individuals from the NBL to this cluster suggests potential challenges in accurately interpreting emotion-triggered muscle weakness. This finding highlights the importance of conducting a comprehensive clinical interview to differentiate cataplexy from its mimics^4^.

Finally, Cluster 1 included predominantly healthy controls, providing a useful baseline for comparison and further emphasizing the phenotypic differences across groups.

The assignment of 8 individuals with a NT1 diagnosis to clusters outside the expected cluster 2 highlights the variability in symptom expression. Cluster 4, with its emphasis on severe fatigue, sleep drunkenness, and functional impairments, included three individuals with definite NT1 and low hypocretin levels, as well as several participants with incomplete diagnostic profiles^35^. These findings underscore the importance of considering atypical presentations in NT1, psychiatric comorbidities, as well as the potential influence of factors such as medication, fluctuating symptoms, or incomplete data (e.g., missing hypocretin-1 or HLA-DQB1 measurements)^36,37^.

Interestingly, the presence of participants with NT1 diagnoses in clusters 3 and 1 suggests the existence of milder or fluctuating phenotypes within the disorder^9^. This could reflect the dynamic nature of symptom expression in NT1, particularly in cases where medication or lifestyle modifications may reduce symptom burden^38–40^. Furthermore, the assignment of four participants from the NBL to the healthy cluster (cluster 1) reinforces the concept that the borderland encompasses a spectrum of severity, with some individuals exhibiting minimal functional impairment^41,42^.

Interestingly, no significant differences were observed in HLA-DQB1 positivity between the two borderland clusters, suggesting that HLA-associated genetic predisposition does not account for the observed disparities. Furthermore, our MSLT analysis showed no significant differences in SOREMPs between the two clusters, aligning with previous findings that REM propensity was not a key differentiating factor^9^. Instead, environmental, behavioral, or other genetic factors may play a more prominent role in shaping the clinical phenotype^43,44^.

Although statistically not significant, data from wearable devices further hinted at potential differences between clusters 3 and 4 in terms of sleep hygiene. Participants in cluster 4 exhibited more variable sleep durations, as well as irregular bedtimes and wake times, indicative of poorer sleep hygiene. This aligns with previous findings, where differences in sleep duration between weekdays and weekends were important for clustering^9^. Such patterns can be indicative of psychiatric comorbidities and may additionally contribute to the greater functional impairment observed in this group. Furthermore, it may represent a modifiable target for interventions aimed at improving daytime functioning^45,46^.

The cluster-based approach highlights phenotypic variability that is not fully captured by current diagnostic criteria for NT1 and narcolepsy borderland^47^. These findings underscore the need for a more dynamic diagnostic framework that incorporates both clinical and biomarker variability^48^. While current criteria rely heavily on features of the multiple sleep latency test, our results suggest that functional impairment and psychiatric comorbidities may also play a critical role in phenotypic differentiation^49,50^. For instance, participants in cluster 4 with a narcolepsy borderland diagnosis exhibited a symptom burden comparable, sometimes even exceeding patients with NT1, highlighting the overlap between these conditions and the potential for misclassification using existing classifications.

It is important to note that prior CDH studies historically have restricted patient inclusion to conform to existing classification criteria (i.e., ICSD3)^9^. The inclusion criteria of the iSPHYNCS observational study is relatively broad which, we feel, better reflects real-world patient populations. Specifically, iSPHYNCS only defines two groups of included participants, i.e., NT1 and healthy controls. All other patients with disorders from the NBL only require a subjective complaint of excessive daytime sleepiness. Patients with comorbid affective disorders may be included, but only if the degree of excessive sleepiness as viewed by the study physician cannot be explained by the comorbid psychiatric condition. The iSPHYNCS approach suggests that psychiatric comorbidities may play an important role in patients who typically present to a sleep disorders clinic. Finally, to what extent Cluster 3 and Cluster 4 may be further divided into additional clusters based on other biomarkers such as microbiome, long-term Fitbit activity, proteomics or other genetic analyses, as are planned for the iSPHYNCS study, remains to be determined. Further work is planned to better understand how patients diagnosed with NT2 or idiopathic hypersomnia based on existing ICSD3 criteria may cluster as the iSPHYNCS multimodal analyses are developed.

This study has several limitations that should be acknowledged. First, while the clustering approach provided meaningful subgroup differentiation, the reliance on self-reported questionnaires introduces the potential for recall bias and subjective variability in symptom reporting. Second, although we included a relatively large sample from multiple centers, the generalizability of our findings may be limited by demographic and clinical differences across study sites. Third, the absence of objective biomarkers such as hypocretin-1 levels for all participants restricts definitive diagnostic confirmation, particularly for individuals with NT1. Additionally, medication effects were not fully controlled, which may have influenced symptom severity and cluster assignment. Furthermore, psychiatric comorbidity data were excluded from the clustering analyses, as only a subset of participant received these questionnaires due to protocol changes, which could have introduced bias based on questionnaire availability rather than participants characteristics. Finally, while our clustering method demonstrated robustness, alternative machine learning approaches could yield different subgroups, warranting further validation in independent cohorts.

### Broader Clinical and Research Implications

The identification of distinct phenotypic clusters has several important implications for clinical practice and research:

1. **Personalized Management**: The variability in symptom burden, psychiatric comorbidities, and sleep hygiene across clusters highlights the need for individualized treatment approaches. For example, participants in cluster 4 may benefit from interventions targeting fatigue and psychiatric symptoms, while those in cluster 3 may require less intensive management.
2. **Refining Diagnostic Understanding**: The overlap of some symptoms between NT1 and narcolepsy borderland, as well as the presence of patients with NT1 in multiple clusters, suggests that current diagnostic criteria may not fully capture clinical variability. Incorporating cluster-based phenotyping could complement traditional diagnostic approaches by highlighting patterns that may inform prognosis or guide individualized care strategies.
3. **Future Research Directions**: Longitudinal studies are needed to assess the stability of cluster assignments over time and to determine whether cluster affiliation predicts clinical outcomes or treatment response. Additionally, further exploration of the underlying mechanisms driving differences between clusters—such as genetic, biological, lifestyle or environmental factors— could yield new insights into the pathophysiology of central disorders of hypersomnolence.

## Conclusions

This study demonstrates the utility of cluster analysis using only questionnaires and easily available metadata in uncovering phenotypic heterogeneity within CDH, challenging traditional diagnostic boundaries and highlighting the need for a more individualized approach to diagnosis and management. By integrating clinical, biomarker, and wearable device data, future research can build on these findings to refine diagnostic frameworks and allow tailored therapies for individuals across the CDH spectrum.

## Supporting information

supplementary sections

## Acknowledgements

We express our gratitude to the participants of the iSPHYNCS study who provided the data for the analyses. A special thanks to the study nurses of the different study centers for their help in recruitment and data collection. We thank the iSPHYNCS Investigators and their teams for their contribution to the realization of this project. We thank Corrado Bernasconi for his advice regarding the statistical analyses.

## Statements

### Ethics Approval

The study was approved by local ethical committees (Ethics Committees ID: 2019– 00788 in Switzerland, 202/2022 in Germany, and NL84710.058.23 in the Netherlands).

### Patient Consent

Each patient was enrolled after providing a written informed consent.

### Permission to Reproduce Material from Other Sources

No material was used from other sources.

### Clinical Trial Registration

The study has been registered in the international clinical trials registry: https://clinicaltrials.gov/study/NCT04330963

## Footnotes

1 [GitHub repository of Missingpy](https://github.com/epsilon-machine/missingpy), *accessed 25.01.2024*

## References

1. Sateia MJ. International classification of sleep disorders. Third edition ed. ICSD. American Academy of Sleep Medicine; 2014.

2. Lammers GJ, Bassetti CLA, Dolenc-Groselj L, et al. Diagnosis of central disorders of hypersomnolence: A reappraisal by European experts. Sleep Med Rev. Aug 2020;52:101306. doi:10.1016/j.smrv.2020.101306

3. Dietmann A, Wenz E, Meer JVd, et al. The Swiss Primary Hypersomnolence and Narcolepsy Cohort Study (SPHYNCS): Study Protocol for a prospective, multicenter cohort observational study. Journal of Sleep Research. 2021;Manuscript accepted for publication.

4. Bassetti CLA, Adamantidis A, Burdakov D, et al. Narcolepsy - clinical spectrum, aetiopathophysiology, diagnosis and treatment. Nat Rev Neurol. Sep 2019;15(9):519–539. doi:10.1038/s41582-019-0226-9

5. Zhang Y, Ren R, Yang L, et al. Comparative polysomnography parameters between narcolepsy type 1/type 2 and idiopathic hypersomnia: a systematic review and meta-analysis. Sleep Medicine Reviews. 2022;63:101610.

6. Zinchuk A, Yaggi HK. Phenotypic subtypes of OSA: a challenge and opportunity for precision medicine. Chest. 2020;157(2):403–420.

7. Venkatnarayan K, Krishnaswamy UM, Rajamuri NKR, et al. Identifying phenotypes of obstructive sleep apnea using cluster analysis. Sleep and Breathing. 2023;27(3):879–886.

8. Bailly S, Destors M, Grillet Y, et al. Obstructive sleep apnea: a cluster analysis at time of diagnosis. PloS one. 2016;11(6):e0157318.

9. Gool JK, Zhang Z, Oei MSSL, et al. Data-Driven Phenotyping of Central Disorders of Hypersomnolence With Unsupervised Clustering. Neurology. 2022;98(23):e2387–e2400. doi:doi:10.1212/WNL.0000000000200519

10. Aellen FM, Van der Meer J, Dietmann A, Schmidt M, Bassetti CLA, Tzovara A. Disentangling the complex landscape of sleep-wake disorders with data-driven phenotyping: A study of the Bernese center. European journal of neurology. Jan 2024;31(1):e16026. doi:10.1111/ene.16026

11. Cook JD, Rumble ME, Plante DT. Identifying subtypes of Hypersomnolence Disorder: a clustering analysis. Sleep Medicine. 2019/12/01/ 2019;64:71-76. 10.1016/j.sleep.2019.06.015

12. Aktan Suzgun M, Wenz ES, van der Meer J, et al. International Swiss Primary Hypersomnolence and Narcolepsy Cohort Study (iSPHYNCS): the impact of psychiatric comorbidities on daily life in central disorders of hypersomnolence—a vicious circle. Journal of Sleep Research. n/a(n/a):e14367. 10.1111/jsr.14367

13. American Academy of Sleep Medicine. International classification of sleep disorders - Third Edition *(ICSD-3)*. Darien, IL: Amercian Academy of Sleep Medicine; 2014.

14. Sheehan DV, Lecrubier Y, Sheehan KH, et al. The Mini-International Neuropsychiatric Interview (MINI): the development and validation of a structured diagnostic psychiatric interview for DSM-IV and ICD-10. Journal of clinical psychiatry. 1998;59(20):22–33.

15. Beck AT, Steer RA, Brown G. Beck depression inventory–II. Psychological assessment. 1996;

16. Zigmond AS, Snaith RP. The hospital anxiety and depression scale. Acta Psychiatr Scand. Jun 1983;67(6):361–70. doi:10.1111/j.1600-0447.1983.tb09716.x

17. Johns MW. A new method for measuring daytime sleepiness: the Epworth sleepiness scale. sleep. 1991;14(6):540–545.

18. Valko PO, Bassetti CL, Bloch KE, Held U, Baumann CR. Validation of the fatigue severity scale in a Swiss cohort. Sleep. Nov 2008;31(11):1601–7. doi:10.1093/sleep/31.11.1601

19. Dauvilliers Y, Beziat S, Pesenti C, et al. Measurement of narcolepsy symptoms: The Narcolepsy Severity Scale. Neurology. Apr 4 2017;88(14):1358–1365. doi:10.1212/WNL.0000000000003787

20. Bargiotas P, Dietmann A, Haynes AG, et al. The Swiss Narcolepsy Scale (SNS) and its short form (sSNS) for the discrimination of narcolepsy in patients with hypersomnolence: a cohort study based on the Bern Sleep-Wake Database. J Neurol. Sep 2019;266(9):2137–2143. doi:10.1007/s00415-019-09365-2

21. Buysse DJ, Reynolds CF, Monk TH, Berman SR, Kupfer DJ. The Pittsburgh Sleep Quality Index: a new instrument for psychiatric practice and research. 28. 1989;28(2):193–213.

22. Dauvilliers Y, Evangelista E, Barateau L, et al. Measurement of symptoms in idiopathic hypersomnia: The Idiopathic Hypersomnia Severity Scale. Neurology. Mar 13 2019;doi:10.1212/WNL.0000000000007264

23. Kanady JC, Harvey AG. Development and Validation of the Sleep Inertia Questionnaire (SIQ) and Assessment of Sleep Inertia in Analogue and Clinical Depression. Cognitive therapy and research. Oct 2015;39(5):601–612. doi:10.1007/s10608-015-9686-4

24. Hays RD, Sherbourne CD, Mazel RM. The RAND 36-Item Health Survey 1.0. Health economics. Oct 1993;2(3):217–27.

25. Weaver TE, Laizner AM, Evans LK, et al. An instrument to measure functional status outcomes for disorders of excessive sleepiness. Sleep. 1997;20(10):835–843.

26. McInnes LH, John; Saul, Nathaniel; Großberger, Lukas. UMAP: Uniform Manifold Approximation and Projection. Journal of Open Source Software. 2018;3(29):861. 10.21105/joss.00861

27. Littner MR, Kushida C, Wise M, et al. Practice parameters for clinical use of the multiple sleep latency test and the maintenance of wakefulness test. Sleep. 2005;28(1):113–121.

28. Gnarra O, van der Meer J, Warncke JD, et al. The Swiss Primary Hypersomnolence and Narcolepsy Cohort Study: feasibility of long-term monitoring with Fitbit smartwatches in central disorders of hypersomnolence and extraction of digital biomarkers in narcolepsy. Sleep. 2024;47(9)doi:10.1093/sleep/zsae083

29. Droogleever Fortuyn HA, Fronczek R, Smitshoek M, et al. Severe fatigue in narcolepsy with cataplexy. J Sleep Res. Apr 2012;21(2):163–9. doi:10.1111/j.1365-2869.2011.00943.x

30. Barateau L, Lopez R, Franchi JAM, Dauvilliers Y. Hypersomnolence, hypersomnia, and mood disorders. Current psychiatry reports. 2017;19:1–11.

31. Vernet C, Leu-Semenescu S, Buzare MA, Arnulf I. Subjective symptoms in idiopathic hypersomnia: beyond excessive sleepiness. Journal of sleep research. 2010;19(4):525–534.

32. Barateau L, Lopez R, Chenini S, et al. Depression and suicidal thoughts in untreated and treated narcolepsy: systematic analysis. Neurology. 2020;95(20):e2755–e2768.

33. Lopez R, Barateau L, Evangelista E, Dauvilliers Y. Depression and hypersomnia: a complex association. Sleep medicine clinics. 2017;12(3):395–405.

34. Zhang Z, Mayer G, Dauvilliers Y, et al. Exploring the clinical features of narcolepsy type 1 versus narcolepsy type 2 from European Narcolepsy Network database with machine learning. Scientific Reports. 2018;8(10628):DOI:10.1038/s41598-018-28840-w.

35. Trotti LM. Waking up is the hardest thing I do all day: Sleep inertia and sleep drunkenness. Sleep Med Rev. Oct 2017;35:76–84. doi:10.1016/j.smrv.2016.08.005

36. Maski K, Steinhart E, Williams D, et al. Listening to the patient voice in narcolepsy: diagnostic delay, disease burden, and treatment efficacy. J Clin Sleep Med. 2017;13(3):419–425.

37. Ozaki A, Inoue Y, Hayashida K, et al. Quality of life in patients with narcolepsy with cataplexy, narcolepsy without cataplexy, and idiopathic hypersomnia without long sleep time: comparison between patients on psychostimulants, drug-naïve patients and the general Japanese population. Sleep medicine. 2012;13(2):200–206.

38. Becker PM, Schwartz JR, Feldman NT, Hughes RJ. Effect of modafinil on fatigue, mood, and health-related quality of life in patients with narcolepsy. Psychopharmacology. 2004;171:133–139.

39. Bogan RK, Black J, Swick T, et al. Correlation of changes in patient-reported quality of life with physician-rated global impression of change in patients with narcolepsy participating in a clinical trial of sodium oxybate: A post hoc analysis. Neurology and therapy. 2017;6:237–245.

40. Emsellem HA, Thorpy MJ, Lammers GJ, et al. Measures of functional outcomes, work productivity, and quality of life from a randomized, phase 3 study of solriamfetol in participants with narcolepsy. Sleep Medicine. 2020;67:128–136.

41. Baumann CR, Mignot E, Lammers GJ, et al. Challenges in diagnosing narcolepsy without cataplexy: a consensus statement. Sleep. Jun 1 2014;37(6):1035–42. doi:10.5665/sleep.3756

42. Nevsimalova S, Susta M, Prihodova I, Horvat EM, Milata M, Sonka K. Idiopathic hypersomnia: a homogeneous or heterogeneous disease? Sleep Med. Apr 2021;80:86–91. doi:10.1016/j.sleep.2021.01.031

43. Ohayon MM. Narcolepsy is complicated by high medical and psychiatric comorbidities: a comparison with the general population. Sleep medicine. 2013;14(6):488–492.

44. Ruoff CM, Reaven NL, Funk SE, et al. High rates of psychiatric comorbidity in narcolepsy: findings from the Burden of Narcolepsy Disease (BOND) study of 9,312 patients in the United States. The Journal of clinical psychiatry. 2017;78(2):19696.

45. Tadrous R, O’Rourke D, Mockler D, Broderick J. Health-related quality of life in narcolepsy: A systematic review and meta-analysis. Journal of Sleep Research. 2021;30(6):e13383. 10.1111/jsr.13383

46. Parmar A, Yeh EA, Korczak DJ, et al. Depressive symptoms, sleep patterns, and physical activity in adolescents with narcolepsy. Sleep. 2019;42(8):zsz111.

47. Fronczek R, Arnulf I, Baumann CR, Maski K, Pizza F, Trotti LM. To split or to lump? Classifying the central disorders of hypersomnolence. Sleep. 2020;43(8):zsaa044.

48. Gauld C, Lopez R, Geoffroy PA, et al. A systematic analysis of ICSD-3 diagnostic criteria and proposal for further structured iteration. Sleep Medicine Reviews. 2021;58:101439.

49. Trotti LM, Staab BA, Rye DB. Test-retest reliability of the multiple sleep latency test in narcolepsy without cataplexy and idiopathic hypersomnia. J Clin Sleep Med. 2013;9(8):789–85.

50. Dietmann A, Gallino C, Wenz E, Mathis J, Bassetti CLA. Multiple sleep latency test and polysomnography in patients with central disorders of hypersomnolence. Sleep Med. Mar 2021;79:6–10. doi:10.1016/j.sleep.2020.12.037

