## supplementary sections for "iSPHYNCS: Unsupervised clustering in questionnaires and metadata reveals distinct subtypes in the narcolepsy borderland"

**iSPHYNCS: Unsupervised clustering in questionnaires and metadata reveals distinct subtypes in the narcoleptic borderland – Supplementary material**

**S1: Questionnaires**

We provide the full set of questions that were used for clustering. The items could be *nominal* (categories without natural order) or *ordinal* (categories with natural order, intervals and ratios). The differentiation between these two types was important for the dimensionality reduction (Sec. S5).

| **Additional Questions** |  |
| --- | --- |
| How often do you remain awake in bed longer than 1 hour in the morning because you lack the energy to get up? | *nom* |
| Have you ever been told or do you yourself suspect that you act out the content of your dreams during sleep? (e.g. hitting, swinging your arms, making running movements, etc.) | *nom* |
| When I want to relax in the evening or sleep at night, do I ever have uncomfortable, restless feelings in my legs that can be relieved by moving or walking around? | *nom* |
| Do your main complaints mainly affect you during the day (unintentional nodding off, tiredness, etc.) or at night (sleep problems, etc.)? | *nom* |
| Of all symptoms you experience, which are the most 1-3 influential on your quality of life? (options: excessive daytime sleepiness, sleep attacks, disturbed nocturnal sleep, not feeling refreshed in the morning, fatigue, non-recreative sleep, difficulty thinking/memorizing/concentrating/staying vigilant, mood swings) | *nom* |
| Are there activities that are important to you, but you cannot perform them (or not to the desired extent) due to your symptoms? (sleep through the night) | *nom* |
| Are there activities that are important to you, but you cannot perform them (or not to the desired extent) due to your symptoms? (perform at school/work how I’d like to) | *nom* |
| Are there activities that are important to you, but you cannot perform them (or not to the desired extent) due to your symptoms? (express emotions freely) | *nom* |
| Are there activities that are important to you, but you cannot perform them (or not to the desired extent) due to your symptoms? (get through the day without sleeping) | *nom* |
| Are there activities that are important to you, but you cannot perform them (or not to the desired extent) due to your symptoms? (sports) | *nom* |
| Are there activities that are important to you, but you cannot perform them (or not to the desired extent) due to your symptoms? (doing chores) | *nom* |
| Are there activities that are important to you, but you cannot perform them (or not to the desired extent) due to your symptoms? (social activities with family and friends) | *nom* |
| Are there activities that are important to you, but you cannot perform them (or not to the desired extent) due to your symptoms? (drive a car or other vehicle (without restrictions) | *nom* |
| Do you have the impression that you have lost work because of your symptoms? | *nom* |
| Do you have the impression that you have difficulties looking after your children because of your symptoms? | *nom* |
| How content are you about your current sleeping habits? | *ord* |
| How worried are you about your current sleep disorder? | *ord* |
| Do you have the impression that you have difficulties concentrating at school/work because of your symptoms? | *ord* |
| Do you have the impression that you do not meet your potential at school/work because of your symptoms? | *ord* |
| Do you have the impression that you have difficulties finding friends at school/work because of your symptoms? | *ord* |
| Do you have the impression that you have been left out at school/work because of your symptoms? | *ord* |
| Do you have the impression that you have difficulties reaching higher positions at school because of your symptoms? | *ord* |
| Do you have the impression that you have limited options in finding a relationship because of your symptoms? | *ord* |
| Do you have the impression that a relationship failed because of your symptoms? | *ord* |
| Do you have the impression that you have difficulties laughing or expressing emotions with your partner and/or children because of your symptoms? | *ord* |
| Do you have the impression that you have difficulties arguing or addressing conflicts with your partner and/or children because of your symptoms? | *ord* |
| Do you have the impression that you have difficulties doing chores because of your symptoms? | *ord* |
| Do you have the impression that you have difficulties going on a vacation because of your symptoms? | *ord* |
| Do you have the impression that you have difficulties going out (e.g., theaters) because of your symptoms? | *ord* |
| Do you have the impression that you have difficulties doing sports because of your symptoms? | *ord* |

| **Complaints Nocturnal Sleep** |  |
| --- | --- |
| Disturbed nocturnal sleep. | *nom* |
| Sleep drunkenness in the morning. | *nom* |
| Sleep drunkenness type. | *nom* |
| Loud (social disturbing) snoring during most nights. | *nom* |
| Interrupted breathing during sleep. | *nom* |
| Violent behaviour during sleep. | *nom* |
| Nightmares. | *nom* |
| Sleep length during the week. | *ord* |
| Sleep length per night during the week. | *ord* |
| How many hours could you theoretically sleep nonstop per night (i.e., when you're on holiday)? | *ord* |
| Estimated subjective sleep latency. | *ord* |
| Habitual time in bed during nighttime sleep episodes on weekdays. | *ord* |
| How many times do you wake up too early in the morning (> 60min before you planned to get up)? | *ord* |
| How often did you sleepwalk as a child? | *ord* |

| **Epworth Sleepiness Scale** |  |
| --- | --- |
| Chance of dozing when sitting and reading | *ord* |
| Chance of dozing when watching TV | *ord* |
| Chance of dozing sitting, inactive in a public place (e.g., a theatre or a meeting) | *ord* |
| Chance of dozing as a passenger in a car for an hour without a break. | *ord* |
| Chance of dozing when lying down to rest in the afternoon when circumstances permit. | *ord* |
| Chance of dozing sitting and talking to someone. | *ord* |
| Chance of dozing when sitting quietly after a lunch without alcohol. | *ord* |
| Chance of dozing in a car while stopped for a few minutes in traffic. | *ord* |

| **Fatigue Severity Scale** |  |
| --- | --- |
| My motivation is lower when I am fatigued. | *ord* |
| Exercise brings on my fatigue. | *ord* |
| I am easily fatigued. | *ord* |
| Fatigue interferes with my physical functioning. | *ord* |
| Fatigue causes frequent problems for me. | *ord* |
| My fatigue prevents sustained physical functioning. | *ord* |
| Fatigue interferes with carrying out certain duties and responsibilities. | *ord* |
| Fatigue is among my three most disabling symptoms. | *ord* |
| Fatigue interferes with my work, family, or social life. | *ord* |

| **Functional Outcome of Sleep Questionnaire** |  |
| --- | --- |
| Do you have difficulty concentrating on the things you do because you are sleepy or tired? | *ord* |
| Do you generally have difficulty remembering things because you are sleepy or tired? | *ord* |
| Do you have difficulty operating a motor vehicle for short durations (less than 1 hour) because you become sleepy or tired? | *ord* |
| Do you have difficulty operating a motor vehicle for long durations (more than 1 hour) because you become sleepy or tired? | *ord* |
| Do you have difficulty visiting with your family or friends in your home because you become sleepy or tired? | *ord* |
| Has your relationship with family, friends or work colleagues been affected because you are sleepy or tired? | *ord* |
| Do you have difficulty watching a movie or videotape because you become sleepy or tired? | *ord* |
| Do you have difficulty getting things done because you are too sleepy or tired to drive or take public transportation? | *ord* |
| Do you have difficulty taking care of financial affairs and doing paperwork because you are sleepy or tired? | *ord* |
| Has your desire for intimacy or sex been affected because you are sleepy or tired? | *ord* |

| **Idiopathic Hypersomnia Severity Scale** |  |
| --- | --- |
| What for you is the ideal duration of night-time sleep (at the weekend or on holiday, for example)? | *ord* |
| When circumstances require that you get up at a particular time in the morning (for example for work or studies, or to take the children to school during the week), do you feel that you have not had enough sleep? | *ord* |
| Is it extremely difficult for you, or even impossible, to wake in the morning without several alarm calls or the help of someone close? | *ord* |
| After a night's sleep, how long does it take you to feel you are functioning properly after you get up (in other words fully functional, both physically and intellectually)? | *ord* |
| In the minutes after waking up, do you ever do irrational things and/or say irrational things, and/or are you very clumsy? | *ord* |
| During the day, when circumstances allow, do you ever take a nap? | *ord* |
| What for you is the ideal length of your naps (at the weekend or on holiday, for example?) Note: if you take several naps, add them all together. | *ord* |
| In general, how do you feel after a nap? | *ord* |
| During the day, while carrying out activities that are not very stimulating, do you ever struggle to stay awake? | *ord* |
| Do you consider that your hypersomnolence has an impact on your general health? | *ord* |
| Do you consider that your hypersomnolence is a problem in terms of your proper intellectual functioning? | *ord* |
| Do you consider that your hypersomnolence affects your mood? | *ord* |
| Do you consider that your hypersomnolence prevents you from carrying out daily tasks properly? | *ord* |
| Do you consider that your hypersomnolence is a problem in terms of your driving a car? | *ord* |

| **Major Complaints** |  |
| --- | --- |
| Do you have complaints about cataplexy? | *nom* |
| Do you have complaints about excessive daytime sleepiness? | *nom* |
| Do you have complaints about hypnagogic hallucinations? | *nom* |
| Do you have complaints about sleep paralysis? | *nom* |

| **Miscellaneous** |  |
| --- | --- |
| What is your gender? | *nom* |
| What is your profession? | *nom* |
| What is your age? | *ord* |
| What is your BMI? | *ord* |

| **Narcolepsy Severity Scale** |  |
| --- | --- |
| Did you experience an irresistible need to sleep during the day? | *ord* |
| Are you worried about falling asleep during the day? | *ord* |
| How important is the disruption of your work/activities caused by these daytime sleep attacks? | *ord* |
| How important is the disruption of your social and family life by these daytime sleep attacks? | *ord* |
| How do you feel generally after one of such daytime sleep attacks? | *ord* |
| After a daytime sleep attack, how much time will pass before the next daytime sleep attack? | *ord* |
| To what extent do these sudden daytime sleep attacks affect your ability to drive a car? | *ord* |
| How frequently do you have episodes of generalized cataplexy when experiencing emotions? | *ord* |
| How frequently do you have episodes of partial cataplexy (only face, neck, arms, or knees) when experiencing emotions? | *ord* |
| How much is your work, social or family life affected by these episodes of cataplexy? | *ord* |
| How frequently do you have hallucinations when falling asleep or waking up? | *ord* |
| To what extent are you bothered by these hallucinations? | *ord* |
| How frequently do you experience sleep paralysis when falling asleep or waking up? | *ord* |
| To what extent are you bothered by these sleep paralysis episodes? | *ord* |
| Currently, how disturbed is your nighttime sleep? | *ord* |

| **Pittsburgh Sleep Quality Index** |  |
| --- | --- |
| During the past month, what time have you usually gone to bed at night? | *ord* |
| During the past month, how long (in minutes) has it usually taken you to fall asleep each night? | *ord* |
| During the past month, what time have you usually gotten up in the morning? | *ord* |
| During the past month, how many hours of actual sleep did you get at night? (This may be different than the number of hours you spent in bed.) | *ord* |
| During the past month, how often have you had trouble sleeping because you cannot get to sleep within 30 minutes? | *ord* |
| During the past month, how often have you had trouble sleeping because you wake up in the middle of the night or early morning? | *ord* |
| During the past month, how often have you had trouble sleeping because you have to get up to use the bathroom? | *ord* |
| During the past month, how often have you had trouble sleeping because you cough or snore loudly? | *ord* |
| During the past month, how often have you had trouble sleeping because you feel too cold? | *ord* |
| During the past month, how often have you had trouble sleeping because you feel too hot? | *ord* |
| During the past month, how often have you had trouble sleeping because you have bad dreams? | *ord* |
| During the past month, how often have you had trouble sleeping because you have pain? | *ord* |
| During the past month, how often have you had trouble sleeping for other reason(s), please describe. | *ord* |
| During the past month, how would you rate your sleep quality overall? | *ord* |
| During the past month, how often have you taken medicine to help you sleep (prescribed or "over the counter")? | *ord* |
| During the past month, how often have you had trouble staying awake while driving, eating meals, or engaging in social activity? | *ord* |
| During the past month, how much of a problem has it been for you to keep up enough enthusiasm to get things done? | *ord* |
| How often in the past month have you had loud snoring? | *ord* |
| How often in the past month have you had long pauses between breaths while asleep? | *ord* |
| How often in the past month have you had legs twitching or jerking while you sleep? | *ord* |
| How often in the past month have you had episodes of disorientation or confusion during sleep? | *ord* |

| **Severity Scores** |  |
| --- | --- |
| Swiss Narcolepsy Scale Score | *ord* |
| Narcolepsy Severity Scale Score | *ord* |
| Fatigue Severity Scale Score | *ord* |
| Epworth Sleepiness Scale Score | *ord* |

| **SF-36** (subset) |  |
| --- | --- |
| During the past 4 weeks, have you had any of the following problems with your work or other regular daily activities as a result of your physical health? Accomplished less than you would like. | *nom* |
| During the past 4 weeks, have you had any of the following problems with your work or other regular daily activities as a result of your physical health? Were limited in the kind of work or other activities. | *nom* |
| During the past 4 weeks, have you had any of the following problems with your work or other regular daily activities as a result of any emotional problems (such as feeling depressed or anxious)? Accomplished less than you would like. | *nom* |
| During the past 4 weeks, have you had any of the following problems with your work or other regular daily activities as a result of any emotional problems (such as feeling depressed or anxious)? Didn't do work or other activities as carefully as usual. | *nom* |
| In general, would you say your health is: | *ord* |
| Does your health limit you in moderate activities? | *ord* |
| The following items are about activities you might do during a typical day. Does your health now limit you in these activities? If so, how much? Climbing several flights of stairs. | *ord* |
| During the past 4 weeks, how much did pain interfere with your normal work (including both work outside the home and housework)? | *ord* |
| These questions are about how you feel and how things have been with you during the past 4 weeks. For each question, please give the one answer that comes closest to the way you have been feeling. Have you felt calm and peaceful? | *ord* |
| These questions are about how you feel and how things have been with you during the past 4 weeks. For each question, please give the one answer that comes closest to the way you have been feeling. Did you have a lot of energy? | *ord* |
| These questions are about how you feel and how things have been with you during the past 4 weeks. For each question, please give the one answer that comes closest to the way you have been feeling. Have you felt downhearted and blue? | *ord* |
| These questions are about how you feel and how things have been with you during the past 4 weeks. For each question, please give the one answer that comes closest to the way you have been feeling. Have you been a happy person? | *ord* |
| During the past 4 weeks, how much of the time has your physical health or emotional problems interfered with your social activities (like visiting with friends, relatives, etc.)? | *ord* |

| **Sleep Inertia Questionnaire** |  |
| --- | --- |
| On a typical morning in the past week, after you wake up, to what extent do you need an alarm to wake up? | *ord* |
| On a typical morning, after you wake up, to what extend do you bump into and drop things? | *ord* |
| On a typical morning, after you wake up, to what extent do you notice that you feel sleepy? | *ord* |
| On a typical morning, after you wake up, to what extent do you notice that it is difficult to keep your balance? | *ord* |
| On a typical morning, after you wake up, to what extent do you feel anxious about the upcoming day? | *ord* |
| On a typical morning, after you wake up, to what extent do you wish you could sleep more? | *ord* |
| On a typical morning, after you wake up, to what extent do you have difficulty concentrating? | *ord* |
| On a typical morning, after you wake up, to what extent do you find that you think more slowly? | *ord* |
| On a typical morning, after you wake up, to what extent do you find that you react more slowly? | *ord* |

| **Swiss Narcolepsy Scale** |  |
| --- | --- |
| How often are you unable to fall asleep? | *ord* |
| How often do you feel bad or not well rested in the morning? | *ord* |
| How often do you take a nap during the day? | *ord* |
| How often have you experienced weak knees/buckling of the knees during emotions like laughing, happiness, or anger? | *ord* |
| How often have you experienced sagging of the jaw during emotions like laughing, happiness, or anger | *ord* |

| **Symptoms Better** |  |
| --- | --- |
| Did you find something that makes your daytime sleepiness better? (Physical activity, sports) | *nom* |
| Did you find something that makes your daytime sleepiness better? (Naps) | *nom* |
| Did you find something that makes your daytime sleepiness better? (Coffee or other stimulants without medication) | *nom* |
| Did you find something that makes your daytime sleepiness better? (Conversation) | *nom* |
| Did you find something that makes your daytime sleepiness better? (Demanding/interesting tasks) | *nom* |
| Did you find something that makes your daytime sleepiness better? (Cold / Fresh air) | *nom* |

| **Symptoms Worse** |  |
| --- | --- |
| Did you find something that makes your daytime sleepiness worse? (Naps) | *nom* |
| Did you find something that makes your daytime sleepiness worse? (Lying down without sleeping) | *nom* |
| Did you find something that makes your daytime sleepiness worse? (Boring tasks) | *nom* |
| Did you find something that makes your daytime sleepiness worse? (Warm surroundings / poor air quality) | *nom* |
| Did you find something that makes your daytime sleepiness worse? (Others) | *nom* |

**S2: Unsupervised Clustering Pipeline**

The pipeline included preprocessing, dimensionality reduction, and consensus clustering.

**Preprocessing**

For the preprocessing, we first encoded categorical data. Ordinal categories were encoded as integer scales, other categories as binary items. Timestamps were transformed into minutes relative to midnight (e.g., 23:55 and 00:05 to -5 and +5, respectively). Then, we excluded individuals if they had answered less than 70% of the questions. In addition, we excluded variables if they were answered by less than 40% of the study participants. We imputed missing data using a random forest imputer. To avoid biasing the imputed values towards the large narcolepsy borderland group, we processed the three cohorts separately (healthy controls, narcolepsy type 1, narcolepsy borderland). For normalization, we scaled each variable to a range from -1 to 1 across all cohorts.

**Dimensionality Reduction**

To give equal weights to all questionnaires during the clustering, we reduced the dimensionality from N dimensions to two dimensions, where N denotes the number of items of each questionnaire.

Since consensus clustering is performed by taking random subsets of the data, we identified the number of clusters and à priori cluster labels for all individuals using all available data. Performing ten repetitions, we applied UMAP for dimensionality reduction (Sec. S3), followed by K-means clustering for k in the range 1 to 11. We identified k = 4 clusters based on the elbow method and the silhouette score (Fig. S1, additional observations in Sec. S5). We set the *à priori* cluster labels of the individuals using consensus clustering based on the labels from the repeated K-means clustering.

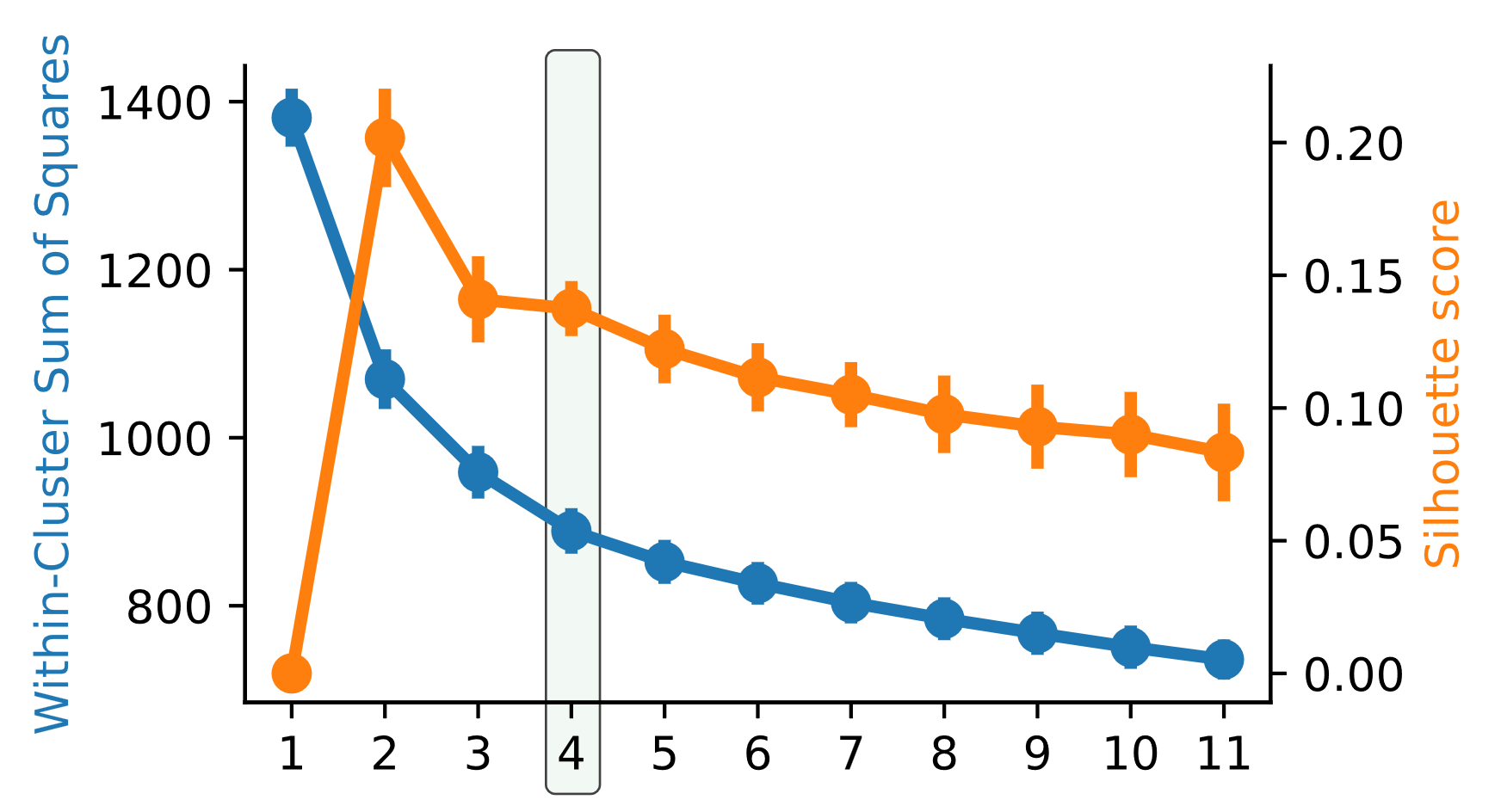

Figure S1. Identification of the number of clusters for k = [1, 11]. At k = 4, the within-cluster sum of squares shows an elbow, and the silhouette score demonstrates a drop. The peak at k = 2 in the silhouette score stems from the separation between healthy controls and the patients with CDH.

To finally obtain the reduced data, we drew 100 random subsets of the data. Each draw included 60% of the individuals stratified by the à priori cluster labels. We applied UMAP to each questionnaire separately to reduce the dimensions from N to 2 in each draw. Finally, each reduction was saved for later used in the conclusive consensus clustering.

**Consensus Clustering**

Using the previously reduced data from 100 repetitions, we applied K-means clustering to the data with k = 4. We calculated the Jaccard coefficient for each repetition to measure how often any pair of individuals happened to be in the same cluster. To set the conclusive label, we applied hierarchical clustering on the resulting Jaccard coefficients to retrieve four clusters.

To assess the goodness of the clustering, we calculated the cluster consensus (Eq. 1) and the individual consensus (Eq. 2), and visualized it in a heatmap (Fig. S2A). To exclude outliers from the clusters, we updated the conclusive cluster label to “undefined” if an individual had an extraordinarily low consensus relative to their cluster (i.e., outlier defined by 3x inter-quartile range, Fig. S2B).

$$m\left( k \right)= \frac{1}{N_{k}(N_{k}-1)/2}\sum_{\begin{aligned} i,j\in I_{k} \\ i<j \end{aligned}} M(i,j)$$

(1)

$$m_{i}\left( k \right)= \frac{1}{N_{k}-1\{e_{i}\in I_{k}\}}\sum_{\begin{aligned} j\in I_{k} \\ j\neq i \end{aligned}} M(i,j)$$

(2)

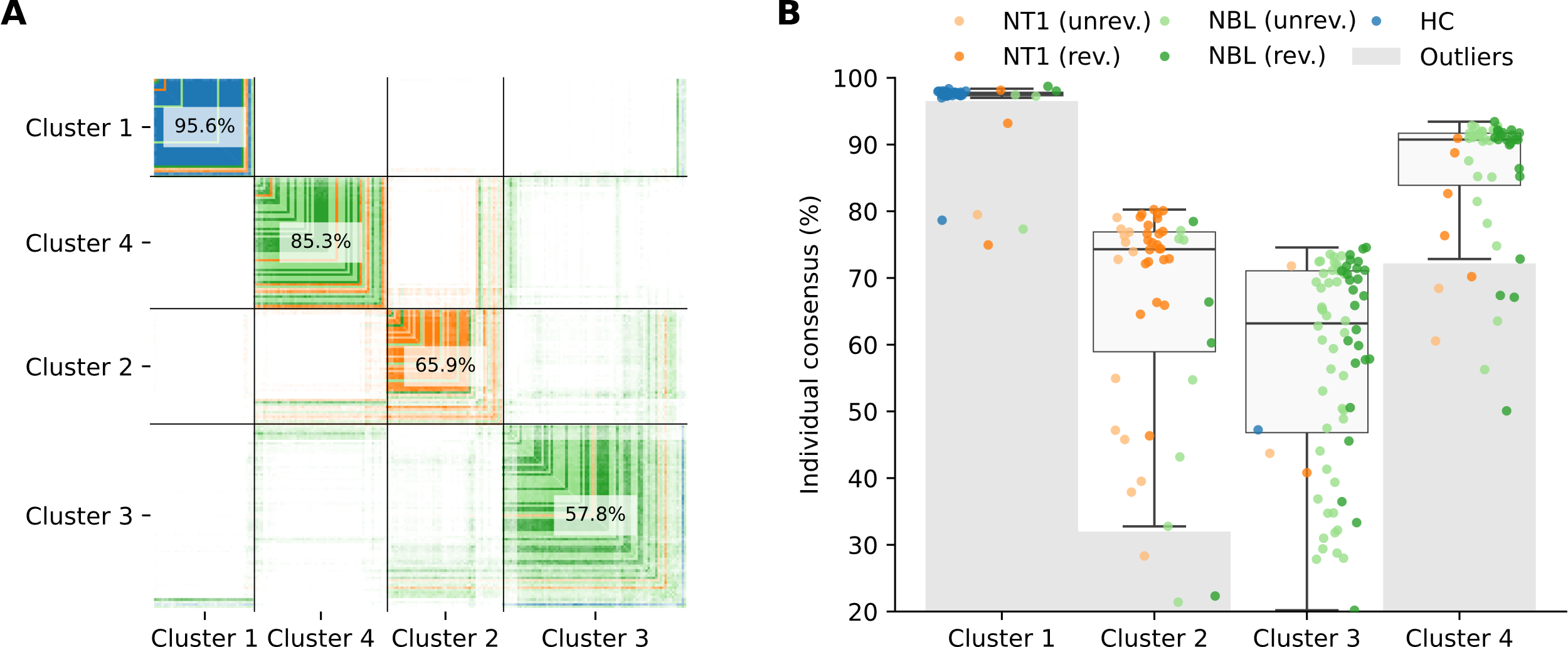

Figure S2. A) Heatmap of the cluster consensus. B) Item consensus by cluster with indicated outliers.

**S3: Dimensionality Reduction of Questionnaires Using UMAP**

Reducing the dimensions for all questionnaires to a common fixed size is beneficial as it gives equal weights to all questionnaires in the clustering algorithm. We tried two approaches experimentally to assess which method is suietd better for the K-means algorithm. Since the K-means algorithm relies on circles or spheres, respectively, we aimed at reductions that best serve this requirement. On the one hand, PCA is unlikely to fulfill this requirement for binary data, although it is appropriate for scales (Fig. S3). UMAP, on the other hand, can fulfill this requirement since it projects the structure of the data into a non-linear representation (Fig. S3). For quality control, we plotted the UMAP projections for all questionnaires using the full dataset for various parameters (Fig. S4). It appeared in all questionnaires that there is a range where perturbations in the parameters would not alter the structure of the projection considerably. Hence, we set the parameters to these ranges for the subsampled UMAP projections depending on the questionnaire and data type. If a questionnaire had mixed data types (i.e., nominal and ordinal), the items were reduced by data type from N to 2 dimensions.

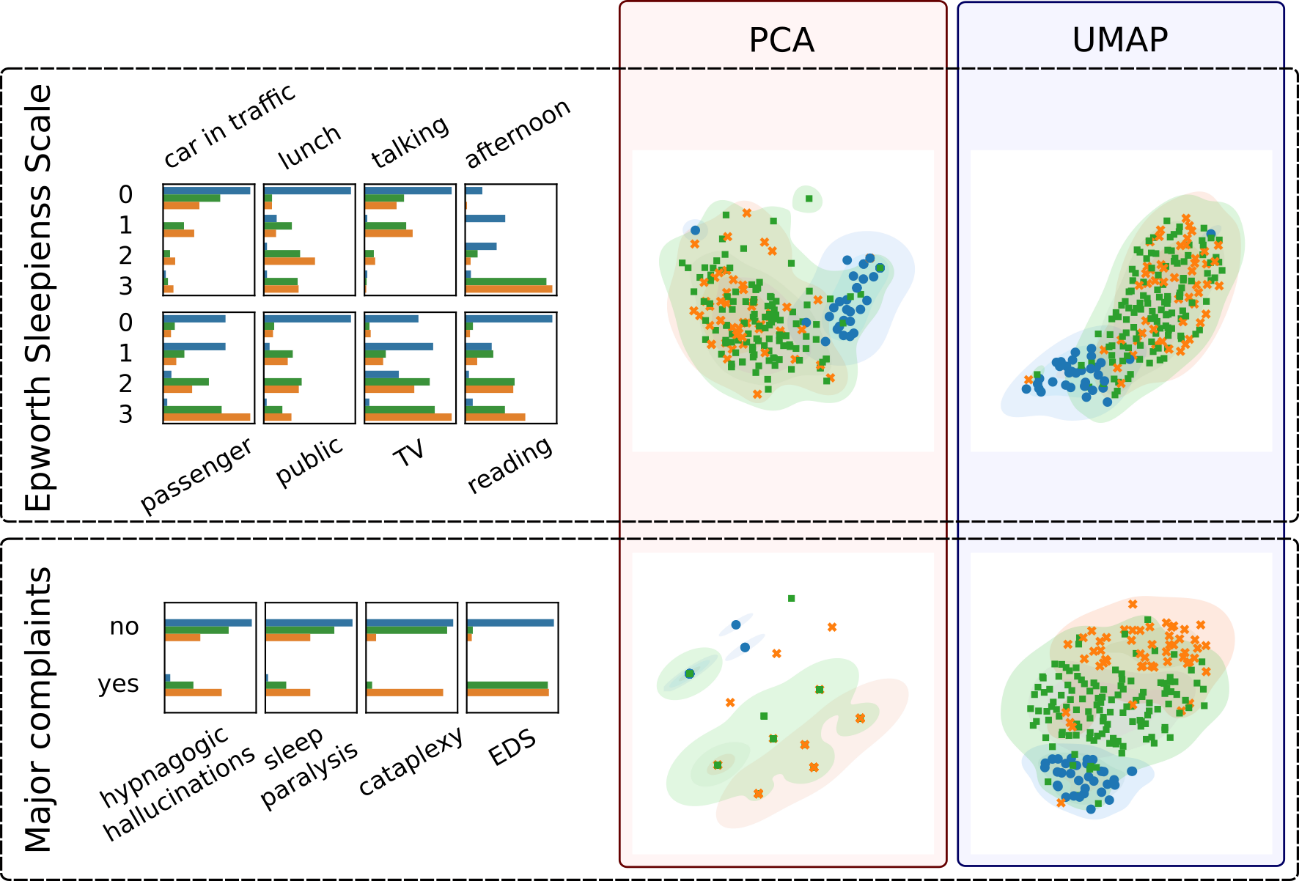

Figure S3: Comparison of two dimensionality reduction algorithms.

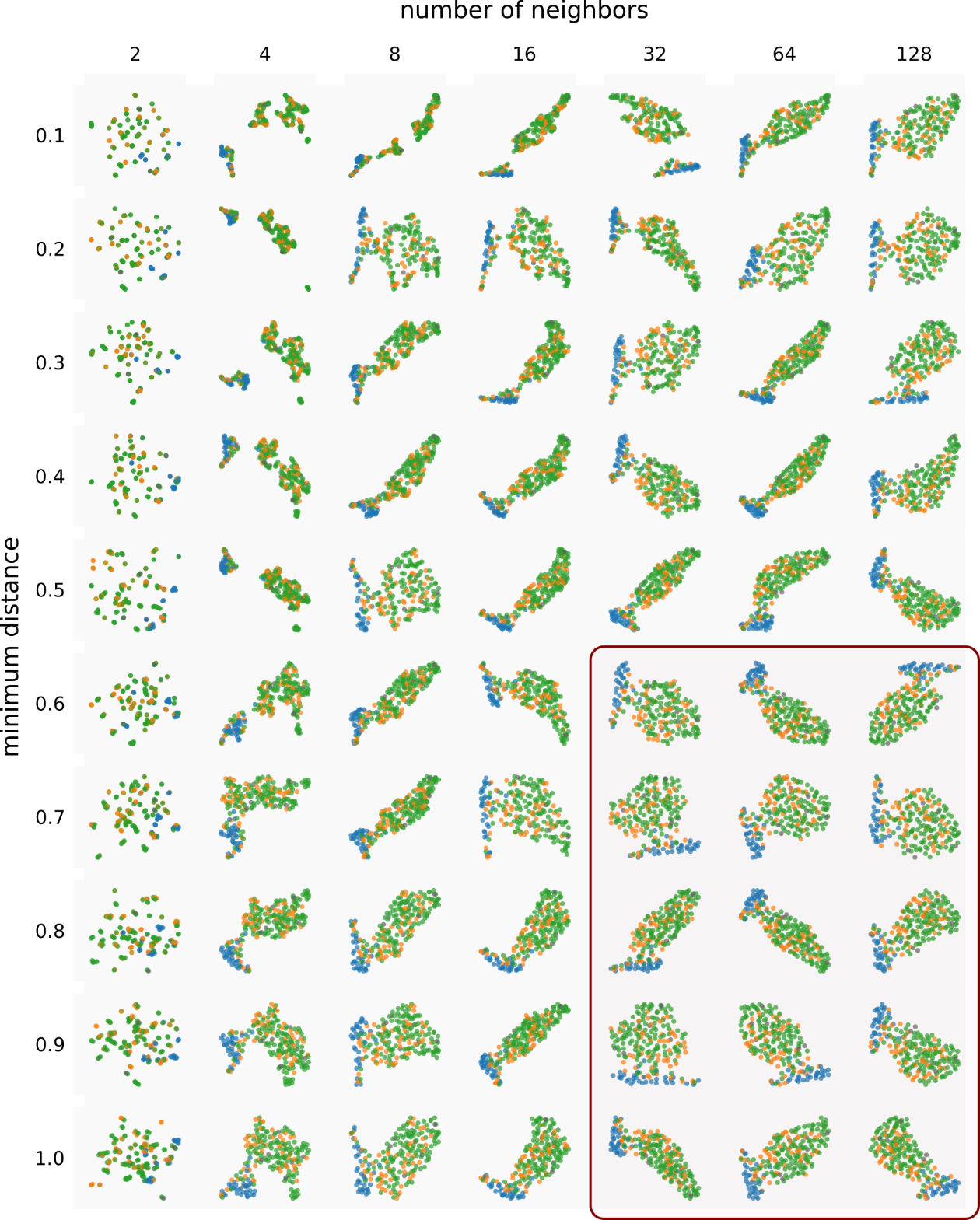

Figure S4. Example of the UMAP projection depending on the number of neighbors and the minimum distance. The structure of the data is stable for larger values (bottom right).

**S4: Most and Least Differentiating Features**

Table S1. Most differentiating variables between cluster 3 and cluster 4. The reported values for the effect size are as follows, depending on the test that was used. **Mann-Whitney U (M-W U)**: Cohen’s d [cluster 3 median (IQR), cluster 4 median (IQR)]; **Ordinal Logistic Regression (OLR)**: Odds Ratio [95% CI]; **Chi-square (C)**: Cramér’s V; **Fisher’s exact (F)**: Odds Ratio.

| **Questionnaire** | **Question** | **Effect size** | **P-value** | **Test** |
| --- | --- | --- | --- | --- |
| Fatigue Severity Scale | Fatigue Severity Scale - Score | **-1.90** [4.67 (4.00, 5.47), 6.50 (5.97, 6.78)] | < 0.001 | M-W U |
| Narcolepsy Severity Scale | Narcolepsy Severity Scale - Score | **-1.33** [11.00 (5.75, 16.25), 22.00 (18.25, 27.00)] | < 0.001 | M-W U |
| Epworth Sleepiness Scale | Epworth Sleepiness Scale - Score | **-1.17** [13.00 (10.00, 15.00), 17.00 (14.00, 19.00)] | < 0.001 | M-W U |
| Sleep Inertia Questionnaire | On a typical morning, after you wake up, to what extent do you have difficulty concentrating? | **18.95** [7.84, 45.82] | < 0.001 | OLR |
| Sleep Inertia Questionnaire | On a typical morning, after you wake up, to what extent do you find that you think react more slowly? | **17.93** [8.05, 39.95] | < 0.001 | OLR |
| Additional Questions | Do you have the impression that you have difficulties concentrating at school/work because of your symptoms? | **17.01** [6.80, 42.56] | < 0.001 | OLR |
| Sleep Inertia Questionnaire | On a typical morning, after you wake up, to what extent do you find that you think more slowly? | **16.15** [7.09, 36.78] | < 0.001 | OLR |
| Idiopathic Hypersomnia Severity Scale | Do you consider that your hypersomnolence is a problem in terms of your proper intellectual functioning? | **16.13** [6.91, 37.64] | < 0.001 | OLR |
| Fatigue Severity Scale | My fatigue prevents sustained physical functioning. | **13.95** [6.53, 29.80] | < 0.001 | OLR |
| SF-36 | Does your health limit you in moderate activities? | **12.01** [5.06, 28.51] | < 0.001 | OLR |
| Epworth Sleepiness Scale | Chance of dozing when lying down to rest in the afternoon when circumstances permit. | **11.19** [1.43, 87.73] | 0.022 | OLR |
| Idiopathic Hypersomnia Severity Scale | Do you consider that your hypersomnolence has an impact on your general health? | **10.65** [4.86, 23.34] | < 0.001 | OLR |
| Pittsburgh Sleep Quality Index | During the past month, how much of a problem has it been for you to keep up enough enthusiasm to get things done? | **10.52** [4.64, 23.88] | < 0.001 | OLR |
| Fatigue Severity Scale | Exercise brings on my fatigue. | **10.42** [5.05, 21.53] | < 0.001 | OLR |
| Idiopathic Hypersomnia Severity Scale | Do you consider that your hypersomnolence prevents you from carrying out daily tasks properly? | **10.02** [4.70, 21.34] | < 0.001 | OLR |
| SF-36 | In general, would you say your health is: | **9.32** [4.31, 20.16] | < 0.001 | OLR |
| Additional Questions | Do you have the impression that you have difficulties doing chores because of your symptoms? | **9.05** [4.31, 18.99] | < 0.001 | OLR |
| Sleep Inertia Questionnaire | On a typical morning, after you wake up, to what extent do you notice that it is difficult to keep your balance? | **9.04** [4.38, 18.66] | < 0.001 | OLR |
| Fatigue Severity Scale | Fatigue interferes with my physical functioning. | **8.60** [4.24, 17.42] | < 0.001 | OLR |
| Fatigue Severity Scale | Fatigue causes frequent problems for me. | **8.16** [3.96, 16.84] | < 0.001 | OLR |
| Fatigue Severity Scale | Fatigue interferes with carrying out certain duties and responsibilities. | **8.10** [4.00, 16.39] | < 0.001 | OLR |
| Narcolepsy Severity Scale | How important is the disruption of your social and family life by these daytime sleep attacks? | **8.02** [3.73, 17.21] | < 0.001 | OLR |
| Fatigue Severity Scale | Fatigue interferes with my work, family, or social life. | **7.78** [3.77, 16.04] | < 0.001 | OLR |
| Narcolepsy Severity Scale | How important is the disruption of your work/activities caused by these daytime sleep attacks? | **7.67** [3.61, 16.30] | < 0.001 | OLR |
| Additional Questions | Do you have the impression that you do not meet your potential at school/work because of your symptoms? | **7.42** [3.54, 15.59] | < 0.001 | OLR |
| Sleep Inertia Questionnaire | On a typical morning, after you wake up, to what extent do you notice that you feel sleepy? | **7.37** [3.26, 16.66] | < 0.001 | OLR |
| Idiopathic Hypersomnia Severity Scale | Do you consider that your hypersomnolence affects your mood? | **7.15** [3.47, 14.76] | < 0.001 | OLR |
| Narcolepsy Severity Scale | How frequently do you have episodes of generalized cataplexy when experiencing emotions? | **6.78** [2.48, 18.54] | < 0.001 | OLR |
| Additional Questions | Do you have the impression that you have difficulties going on a vacation because of your symptoms? | **6.50** [3.18, 13.27] | < 0.001 | OLR |
| Fatigue Severity Scale | I am easily fatigued. | **6.01** [3.01, 12.00] | < 0.001 | OLR |
| Additional Questions | Do you have the impression that you have limited options in finding a relationship because of your symptoms? | **5.93** [2.91, 12.09] | < 0.001 | OLR |
| Additional Questions | Do you have the impression that you have difficulties doing sports because of your symptoms? | **5.72** [2.85, 11.45] | < 0.001 | OLR |
| SF-36 | These questions are about how you feel and how things have been with you during the past 4 weeks. For each question, please give the one answer that comes closest to the way you have been feeling. Did you have a lot of energy? | **5.55** [2.72, 11.29] | < 0.001 | OLR |
| Narcolepsy Severity Scale | How do you feel generally after one of such daytime sleep attacks? | **5.53** [2.64, 11.56] | < 0.001 | OLR |
| Sleep Inertia Questionnaire | On a typical morning, after you wake up, to what extent do you bump into and drop things? | **5.17** [2.62, 10.20] | < 0.001 | OLR |
| Idiopathic Hypersomnia Severity Scale | During the day, while carrying out activities that are not very stimulating, do you ever struggle to stay awake? | **5.09** [2.53, 10.27] | < 0.001 | OLR |
| Sleep Inertia Questionnaire | On a typical morning, after you wake up, to what extent do you feel anxious about the upcoming day? | **5.08** [2.56, 10.09] | < 0.001 | OLR |
| Fatigue Severity Scale | Fatigue is among my three most disabling symptoms. | **5.05** [2.41, 10.59] | < 0.001 | OLR |
| Idiopathic Hypersomnia Severity Scale | In the minutes after waking up, do you ever do irrational things and/or say irrational things, and/or are you very clumsy? | **4.99** [2.42, 10.28] | < 0.001 | OLR |
| Narcolepsy Severity Scale | Did you experience an irresistible need to sleep during the day? | **4.78** [2.41, 9.47] | < 0.001 | OLR |
| Additional Questions | Do you have the impression that you have difficulties laughing or expressing emotions with your partner and/or children because of your symptoms? | **4.75** [2.38, 9.48] | < 0.001 | OLR |
| Idiopathic Hypersomnia Severity Scale | When circumstances require that you get up at a particular time in the morning (for example, for work or studies, or to take the children to school during the week), do you feel that you have not had enough sleep? | **4.66** [1.78, 12.22] | 0.002 | OLR |
| Idiopathic Hypersomnia Severity Scale | What for you is the ideal length of your naps (at the weekend or on holiday, for example?) Note: if you take several naps, add them all together. | **4.45** [2.17, 9.10] | < 0.001 | OLR |
| Epworth Sleepiness Scale | Chance of dozing sitting and talking to someone. | **4.45** [2.12, 9.32] | < 0.001 | OLR |
| Swiss Narcolepsy Scale | How often have you experienced sagging of the jaw during emotions like laughing, happiness, or anger | **4.44** [1.57, 12.58] | 0.005 | OLR |
| SF-36 | The following items are about activities you might do during a typical day. Does your health now limit you in these activities? If so, how much? Climbing several flights of stairs. | **4.44** [2.11, 9.34] | < 0.001 | OLR |
| Additional Questions | Do you have the impression that you have difficulties arguing or addressing conflicts with your partner and/or children because of your symptoms? | **4.32** [2.22, 8.42] | < 0.001 | OLR |
| Narcolepsy Severity Scale | After a daytime sleep attack, how much time will pass before the next daytime sleep attack? | **4.30** [2.18, 8.46] | < 0.001 | OLR |
| Epworth Sleepiness Scale | Chance of dozing sitting, inactive in a public place (e.g., a theatre or a meeting) | 4**.26** [2.09, 8.67] | < 0.001 | OLR |
| Additional Questions | Do you have the impression that you have difficulties reaching higher positions at school because of your symptoms? | **4.22** [2.16, 8.24] | < 0.001 | OLR |
| Additional Questions | Do you have the impression that a relationship failed because of your symptoms? | **4.20** [2.00, 8.84] | < 0.001 | OLR |
| Pittsburgh Sleep Quality Index | During the past month, how often have you had trouble staying awake while driving, eating meals, or engaging in social activity? | **4.05** [2.03, 8.08] | < 0.001 | OLR |
| Fatigue Severity Scale | My motivation is lower when I am fatigued. | **4.04** [2.00, 8.17] | < 0.001 | OLR |
| Functional Outcome of Sleep Questionnaire | Do you have difficulty getting things done because you are too sleepy on tired to drive or take public transportation? | **0.20** [0.10, 0.40] | < 0.001 | OLR |
| Functional Outcome of Sleep Questionnaire | Do you have difficulty taking care of financial affairs and doing paperwork because you are sleepy or tired? | **0.17** [0.08, 0.37] | < 0.001 | OLR |
| SF-36 | During the past 4 weeks, how much of the time has your physical health or emotional problems interfered with your social activities (like visiting with friends, relatives, etc.)? | **0.16** [0.08, 0.33] | < 0.001 | OLR |
| Functional Outcome of Sleep Questionnaire | Do you have difficulty visiting with your family or friends in your home because you become sleepy or tired? | **0.12** [0.06, 0.25] | < 0.001 | OLR |
| Functional Outcome of Sleep Questionnaire | Has your relationship with family, friends or work colleagues been affected because you are sleepy or tired? | **0.11** [0.05, 0.23] | < 0.001 | OLR |
| Functional Outcome of Sleep Questionnaire | Do you generally have difficulty remembering things because you are sleepy or tired? | **0.09** [0.04, 0.19] | < 0.001 | OLR |
| Functional Outcome of Sleep Questionnaire | Do you have difficulty concentrating on the things you do because you are sleepy or tired? | **0.06** [0.03, 0.15] | < 0.001 | OLR |
| SF-36 | During the past 4 weeks, have you had any of the following problems with your work or other regular daily activities as a result of your physical health? Were limited in the kind of work or other activities. | **13.43** | < 0.001 | F |
| Additional Questions | Are there activities that are important to you, but you cannot perform them (or not to the desired extent) due to your symptoms? (option: perform at school/work how I would like to) | **11.26** | < 0.001 | F |

Table S2. Least differentiating variables between cluster 3 and cluster 4. The reported values for the effect size are as follows, depending on the test that was used. **Mann-Whitney U (M-W U)**: Cohen’s d [cluster 3 median (IQR), cluster 4 median (IQR)]; **Ordinal Logistic Regression (OLR)**: Odds Ratio [95% CI]; **Chi-square (C)**: Cramér’s V; **Fisher’s exact (F)**: Odds Ratio.

| **Questionnaire** | **Question** | **Effect size** | **P-value** | **Test** |
| --- | --- | --- | --- | --- |
| Pittsburgh Sleep Quality Index | During the past month, what time have you usually gotten up in the morning? | **0.19** [405.00 (375.00, 479.00), 420.00 (360.00, 476.25)] | 0.858 | M-W U |
| Pittsburgh Sleep Quality Index | During the past month, how long (in minutes) has it usually taken you to fall asleep each night? | **0.11** [10.00 (5.00, 20.00), 5.00 (5.00, 13.75)] | 0.018 | M-W U |
| Pittsburgh Sleep Quality Index | During the past month, what time have you usually gone to bed at night? | **0.08** [-90.00 (-120.00, -52.50), -90.00 (-120.00, -60.00)] | 0.749 | M-W U |
| Complaints Nocturnal Sleep | Habitual time in bed during nighttime sleep episodes on weekdays [Hours]. | **0.06** [8.50 (8.00, 9.50), 9.00 (8.00, 10.00)] | 0.677 | M-W U |
| Complaints Nocturnal Sleep | Estimated subjective sleep latency. | **-0.02** [10.00 (5.00, 15.00), 5.00 (3.00, 15.00)] | 0.169 | M-W U |
| Miscellaneous | What is your BMI? | **-0.03** [23.30 (20.35, 26.13), 23.60 (21.20, 30.10)] | 0.093 | M-W U |
| Pittsburgh Sleep Quality Index | During the past month, how often have you had trouble sleeping for other reason(s), please describe. | **1.42** [0.61, 3.28] | 0.415 | OLR |
| Pittsburgh Sleep Quality Index | During the past month, how often have you had trouble sleeping because you feel too cold? | **1.37** [0.68, 2.75] | 0.383 | OLR |
| Pittsburgh Sleep Quality Index | During the past month, how often have you had trouble sleeping because you have to get up to use the bathroom? | **1.32** [0.68, 2.59] | 0.411 | OLR |
| Pittsburgh Sleep Quality Index | During the past month, how often have you had trouble sleeping because you cannot get to sleep within 30 minutes? | **1.27** [0.65, 2.47] | 0.488 | OLR |
| Pittsburgh Sleep Quality Index | During the past month, how often have you had trouble sleeping because you feel too hot? | **1.23** [0.61, 2.47] | 0.557 | OLR |
| Pittsburgh Sleep Quality Index | During the past month, how often have you had trouble sleeping because you have bad dreams? | **1.21** [0.61, 2.42] | 0.584 | OLR |
| Additional Questions | How worried are you about your current sleep disorder? | **1.00** [0.62, 1.62] | 0.998 | OLR |
| Additional Questions | How content are you about your current sleeping habits? | **1.00** [0.63, 1.59] | 0.999 | OLR |
| Swiss Narcolepsy Scale | How often are you unable to fall asleep? | **0.99** [0.52, 1.90] | 0.988 | OLR |
| Pittsburgh Sleep Quality Index | During the past month, how often have you had trouble sleeping because you wake up in the middle of the night or early morning? | **0.98** [0.51, 1.89] | 0.955 | OLR |
| Pittsburgh Sleep Quality Index | During the past month, how often have you had trouble sleeping because you cough or snore loudly? | **0.95** [0.36, 2.49] | 0.922 | OLR |
| Complaints Nocturnal Sleep | How often did you sleep walk as a child? | **0.81** [0.39, 1.68] | 0.571 | OLR |
| Complaints Hypnagogic Hallucinations | Do you have complaints about hypnagogic hallucinations? | **1.9** | 0.114 | F |
| Symptoms Worse | Did you find something that makes your daytime sleepiness worse? (Demanding/interesting tasks) | **1.67** | 0.48 | F |
| Additional Questions | Of all symptoms you experience, which are the most 1-3 influential on your quality of life? (option: lightheadedness) | **1.66** | 0.674 | F |
| Additional Questions | Of all symptoms you experience, which are the most 1-3 influential on your quality of life? (option: hypnagogic hallucinations) | **1.63** | 1 | F |
| Symptoms Worse | Did you find something that makes your daytime sleepiness worse? (Smoking) | **1.63** | 1 | F |
| Additional Questions | Of all symptoms you experience, which are the most 1-3 influential on your quality of life? (option: fatigue) | **1.62** | 0.373 | F |
| Additional Questions | Of all symptoms you experience, which are the most 1-3 influential on your quality of life? (option: excessive daytime sleepiness) | **1.53** | 0.271 | F |
| Complaints Nocturnal Sleep | Disturbed nocturnal sleep. | **1.48** | 0.402 | F |
| Symptoms Better | Did you find something that makes your daytime sleepiness better?(Others) | **1.39** | 0.747 | F |
| Symptoms Better | Did you find something that makes your daytime sleepiness better?(Cold / Fresh air) | **1.35** | 0.545 | F |
| Symptoms Worse | Did you find something that makes your daytime sleepiness worse? (Warm surroundings / poor air quality) | **1.34** | 0.451 | F |
| Complaints Sleep Paralysis | Do you have complaints about sleep paralysis? | **1.32** | 0.524 | F |
| Miscellaneous | What is your gender? | **1.24** | 0.655 | F |
| Symptoms Worse | Did you find something that makes your daytime sleepiness worse?(Lying down without sleeping) | **1.12** | 0.844 | F |
| Additional Questions | Of all symptoms you experience, which are the most 1-3 influential on your quality of life? (option: excessive weight gain) | **1.09** | 1 | F |
| Additional Questions | Are there activities that are important to you, but you cannot perform them (or not to the desired extent) due to your symptoms? (option: others) | **1.08** | 1 | F |
| Symptoms Worse | Did you find something that makes your daytime sleepiness worse? (Others) | **0.92** | 1 | F |
| Complaints Nocturnal Sleep | Do you have complaints about sleep walking? | **0.91** | 1 | F |
| Additional Questions | Of all symptoms you experience, which are the most 1-3 influential on your quality of life? (option: hyperactivity) | **0.8** | 1 | F |
| Symptoms Better | Did you find something that makes your daytime sleepiness better? (Naps) | **0.79** | 0.682 | F |
| Symptoms Better | Did you find something that makes your daytime sleepiness better? (Demanding/interesting tasks) | **0.77** | 0.559 | F |
| Symptoms Better | Did you find something that makes your daytime sleepiness better? (Physical activity, sports) | **0.69** | 0.357 | F |
| Additional Questions | Of all symptoms you experience, which are the most 1-3 influential on your quality of life? (option: sleep attacks) | **0.67** | 0.74 | F |
| Symptoms Better | Did you find something that makes your daytime sleepiness better? (Coffee or other stimulants without medication) | **0.66** | 0.497 | F |
| Complaints Nocturnal Sleep | Violent behaviour during sleep. | **0.03** | 0.956 | C |

**S5: Clustering with different numbers of clusters**

Previously, we identified the ideal number of clusters to be k = 4 (Sec. S2). In addition to running the analyses for that number, we also provide the consensus charts and Sankey plots for k = 3 and k = 5 (Fig. S5).

For k = 3, the majority of patients with NT1 fall in the same cluster, with roughly half the patients in the NBL. Although this is not problematic per se, we appreciate the presence of a dedicated cluster for patients with NT1 at k = 4 as an indication for successful clustering, since this was expected. For k = 5, there is a cluster containing only n = 4 individuals. In addition, the cluster consensus dropped substantially in all clusters except for cluster 1.

With these observations on top of the previously used metrics (within-sum of squares and silhouette score), we conclude that k = 4 is likely the best number of clusters given the observed data.

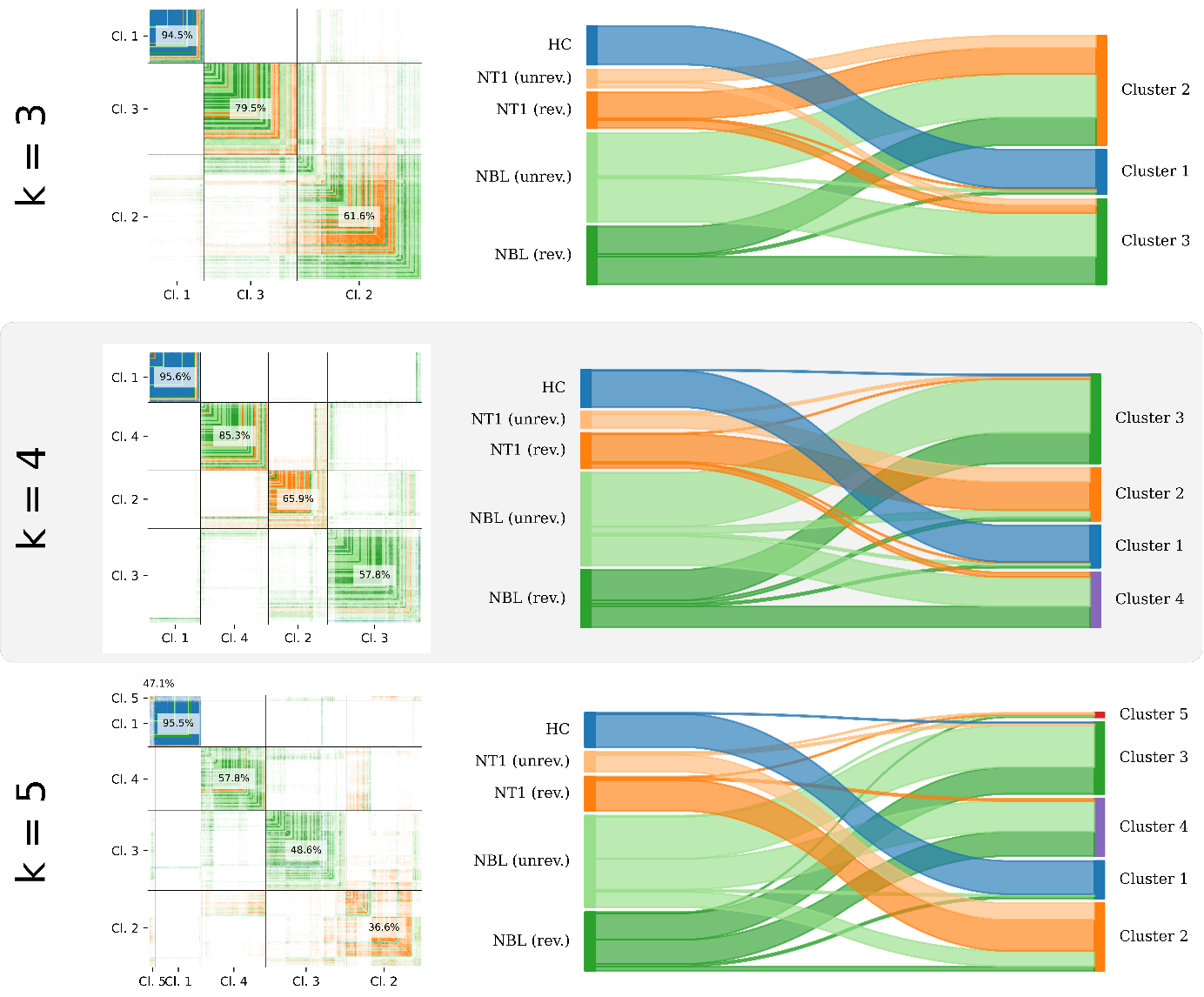

Figure S5. Consensus chart and Sankey plot for k = {3, 4, 5}.
